# Scalable Epidemiological Workflows to Support COVID-19 Planning and Response

**DOI:** 10.1101/2021.02.23.21252325

**Authors:** Dustin Machi, Parantapa Bhattacharya, Stefan Hoops, Jiangzhuo Chen, Henning Mortveit, Srinivasan Venkatramanan, Bryan Lewis, Mandy Wilson, Arindam Fadikar, Tom Maiden, Christopher L. Barrett, Madhav V. Marathe

## Abstract

The COVID-19 global outbreak represents the most significant epidemic event since the 1918 influenza pandemic. Simulations have played a crucial role in supporting COVID-19 planning and response efforts. Developing scalable workflows to provide policymakers quick responses to important questions pertaining to logistics, resource allocation, epidemic forecasts and intervention analysis remains a challenging computational problem. In this work, we present scalable high performance computing-enabled workflows for COVID-19 pandemic planning and response. The scalability of our methodology allows us to run fine-grained simulations daily, and to generate county-level forecasts and other counter-factual analysis for each of the 50 states (and DC), 3140 counties across the USA. Our workflows use a hybrid cloud/cluster system utilizing a combination of local and remote cluster computing facilities, and using over 20,000 CPU cores running for 6–9 hours every day to meet this objective. Our state (Virginia), state hospital network, our university, the DOD and the CDC use our models to guide their COVID-19 planning and response efforts. We began executing these pipelines March 25, 2020, and have delivered and briefed weekly updates to these stakeholders for over 30 weeks without interruption.

## I. Introduction

COVID-19 represents the first pandemic since the 2009 H1N1 outbreak and is the worst pandemic on record since the 1918 pandemic. Since February 2020, the pandemic has had a severe economic, social, and health impact. According to the International Monetary Fund (IMF), the global economic burden for COVID-19 will likely be 9+ trillion US dollars. More than 30 million confirmed infections and 1 million deaths have been reported globally, with very different epidemic dynamic trajectories and mortality witnessed across various countries. Europe and the United States (US) are seeing a resurgence of cases and the situation is unlikely to get better anytime soon.

Epidemiological models and workflows comprising of these models can help provide insight into the spatiotemporal dynamics of epidemics by: (*i*) forecasting the epidemic’s future course, (*ii*) guiding allocation of scarce resources and assessing depletion of current resources, (*iii*) inferring disease parameters that allow researchers to make better recommendations and (*iv*) providing insight into the effectiveness of different interventions. Individual behavior and public policies are critical influencers for controlling epidemics, and computational simulations can be powerful tools for understanding which behaviors and policies are likely to be effective. Our studies have used meta-population models, as well as detailed agent-based models. The network-based models consider epidemic spread on an undirected social interaction network *G*(*V, E*) over a population *V*, where each edge *e* = (*u, v*) ∈ *E* implies that individuals (also referred to as nodes) *u, v* ∈ *V* interact [16], [34] ^1^.

### Our contributions and significance

In this paper, we describe a novel high performance computing (HPC) approach for executing epidemiological workflows that can support planning and response to pandemics such as COVID-19. Our approach is unique: (*i*) it uses detailed agent-based models as well as meta-population models to simulate epidemic dynamics over realistic representations of national-scale social contact networks, (*ii*) it splits the workflow across two supercomputing clusters due to resource constraints, and (*iii*) it is used to support near real-time response efforts. The workflows are comprised of a complex series of data ingestion, simulation and analytics steps. Details of how EpiHiper, the agent-based discrete time simulator for infectious disease spread used in this work, and other such networked agent-based modeling frameworks work are described in companion publications and are not the focus of this paper. However, the basic approach presented here can be used for other agent models and other synthetic social contact networks. We focus here on three epidemic workflows: (*i*) calibration of the models using county-level incidence data, (*ii*) predicting daily county-level incidence values for time periods covering two weeks to a few months and (*iii*) counter-factual analysis of various policy decisions during the ongoing pandemic. Key steps in all of the workflows include (*i*) a data-driven algorithm that integrates county-level incidence data, as well as individual behavioral representations and public policies, to calibrate the models and project incidence going forward; (*ii*) realistic individual-level social contact networks and HPC agent-based models to produce highly resolved outcomes (at the individual and family levels); and (*iii*) analytics that combine the simulation output, surveillance data and detailed synthetic data to support policy assessment. The workflows are executed in real time, meaning that the pipeline produces epidemic predictions every week that we share with federal and state authorities. Splitting and orchestration of workflows across two supercomputing systems that are geographically separated requires careful analysis of the workload and practical constraints.

We demonstrate our results by showing how our epidemiological workflows can be used to support a national COVID-19 response. Our pipeline typically runs 5,000– 17,900 simulations per night, covering the entire US network which is comprised of about 300 million nodes and 7.9 billion edges partitioned across all 50 states and Washington DC. The simulations yield ensemble models for prediction of epidemic incidence curves at the US county level (3140 counties). These results are the first of their kind reported in the literature for national-scale US networks. The workflow is orchestrated between home and remote super-computing clusters; 20,000 cores of the remote super-computing cluster are dedicated each night for completing our complex calibration and prediction tasks.

We are the lead modeling group supporting our state’s (Virginia) COVID-19 response. We have provided uninterrupted weekly projections and analytical products to the analysts and senior officials of the state hospital referral regions (HRR) and local universities (including our university) since March 25, 2020.^2^. We also provide our weekly forecasts to the Centers for Disease Control and Prevention (CDC), and our analytical products to the Department of Defense (DoD). Our results demonstrate that *real-time, data-driven high resolution epidemics science at a national scale is indeed possible*^3^.

### Overview

The epidemiological pipeline workflows are shown in Figures 1, 3, 5 and 4. Each workflow is split between a local cluster (Rivanna HPC Facility at University of Virginia) and a remote super-computing cluster (Bridges HPC Facility at Pittsburgh Supercomputing Center). Our workflows support our vision of *HPC-oriented, real-time epidemic science*, and are carefully organized to conform to the following set of practical constraints. (*i*) Our access to the remote super-computing cluster is limited, so it is not available to us 24/7. We note that the level of access provided to us is very generous — we have had exclusive access to the cluster, with over 20,000 cores, for 10 hours a day (from 10 pm to 8 am) for over 4 months. (*ii*) Purchased datasets and tools are maintained on our home cluster; these items are not ported to the remote super-computing cluster due to time and licensing requirements. (*iii*) Analysts have more consistent access to and control over the home cluster, so the workflows are split between the two sites. The workflows are also designed to provide a level of resiliency and task parallelization: we use the local cluster during the day, and use the remote super-computing cluster at night. Figure 2 shows the timeline of tasks involving human efforts. The overall workflow differs based on the specific kind of problem addressed, but all of them consist of the following significant sub-components: (*i*) generation of national-scale synthetic social contact networks, (*ii*) agent-based and meta-population models that can scale to large systems, (*iii*) methods for calibrating and producing ensemble models, (*iv*) tools for assembling the input data and distributing this dataset on cluster nodes, and (*v*) tools for post-processing the output data so that summary data can be sent back to the home cluster for further analysis.

**Figure 1.**
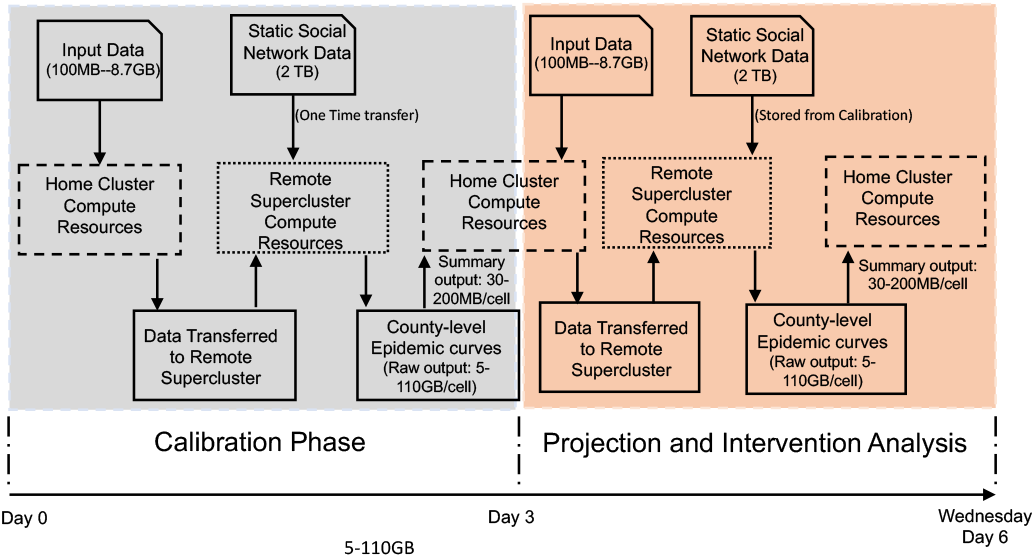
Combined workflow: This diagram illustrates the complete timeline of our process, from model configuration through intervention analysis.

**Figure 2.**
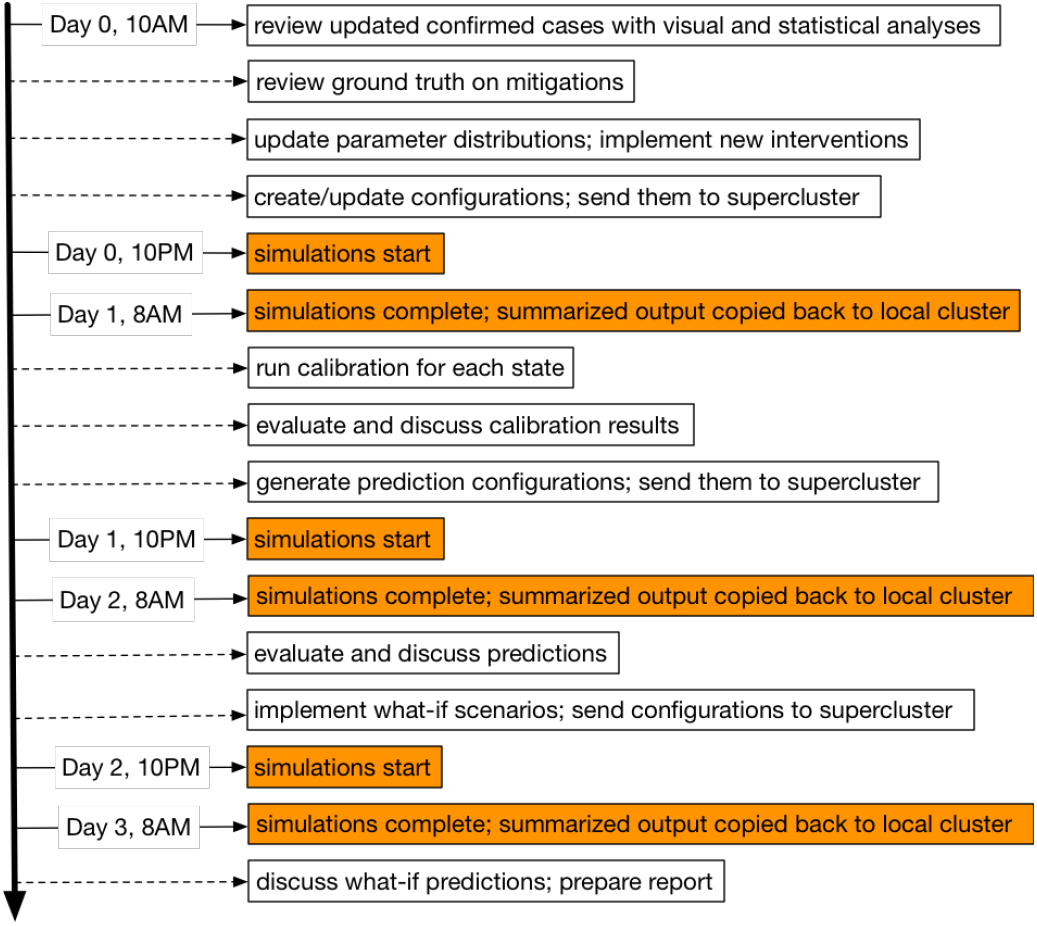
Timeline of tasks involving human efforts. It shows the schedule of the sequence of tasks over multiple days for a complete calibration-prediction cycle. The orange boxes are automated.

**Figure 4.**
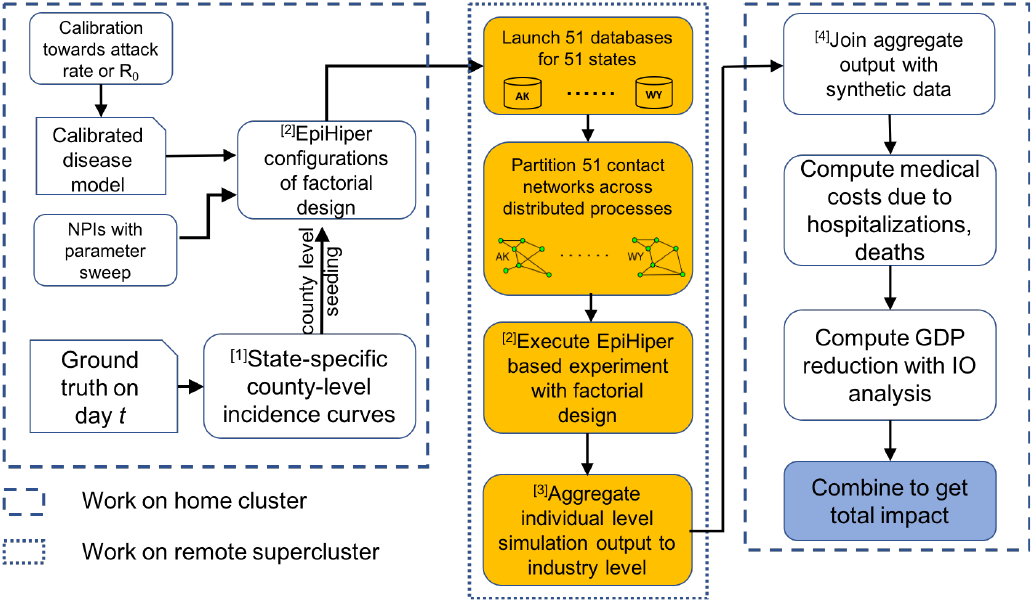
Calibration workflow: [1] Incidence data includes about 3000 counties × over 200 days of entries. [2] An example of calibration design has 300 cells × 51 states × 1 replicates = 15300 simulation instances. [3] Size of individual level output data: 300 cells × 51 states × 1 replicates × multi-million state transitions = multi-billion entries, about 5TB. [4] Calibration uses aggregate data of size: 300 cells × 51 states × 1 replicates × 365 days × 90 health states × 3 counts = about 1.5 billion entries, 4GB.

**Figure 3.**
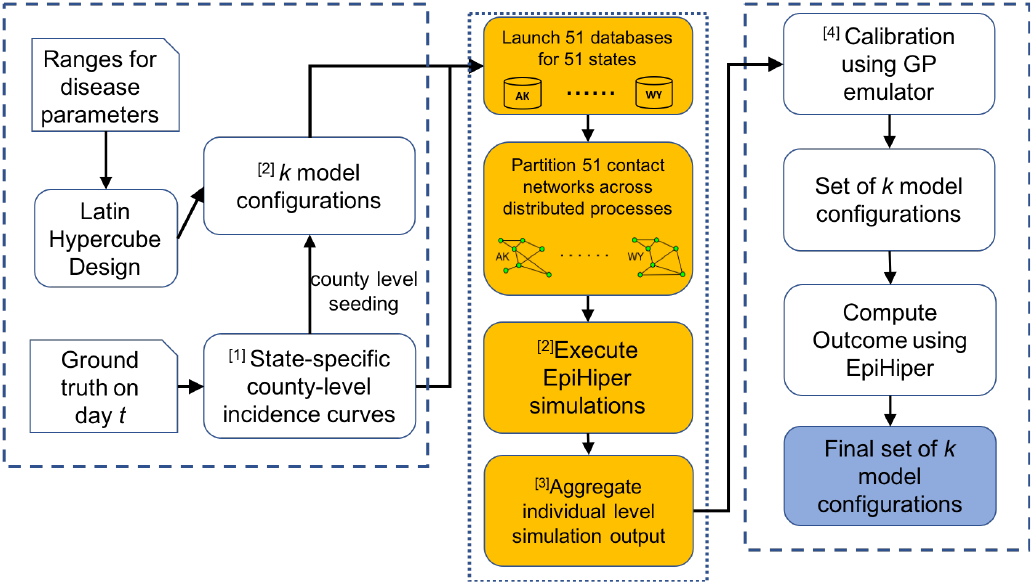
Economic workflow: Economic workflow is used for computing the medical costs incurred due to the pandemic. The models and details of this can be found in [9]. [1] Incidence data includes about 3000 counties× over 200 days of entries. [2] An example factorial design has (2 VHI compliances × 3 lockdown durations × 2 lockdown compliances) × 51 states × 15 replicates = 9180 simulation instances. [3] Size of individual level output data: 12 cells× 51 states × 15 replicates × multi-million state transitions = multi-billion entries, about 3TB. [4] Size of aggregate output data: 12 cells × 51 states × 15 replicates × 365 days × 90 health states × 3 counts = about 1 billion entries, 2.5GB. Size of synthetic data: 300 million × 8 = 2.5 billion entries.

**Figure 5.**
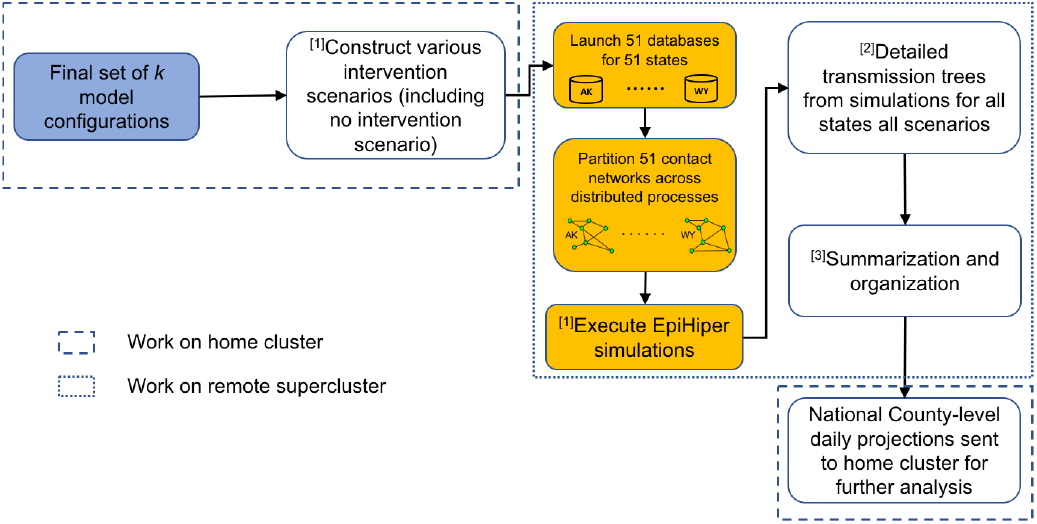
Prediction workflow: [1] An example design has (3 partial reopening levels × 4 contact tracing compliances) × 51 states × 15 replicates = 9180 simulation instances. [2] Size of transmission tree data: 12 cells× 51 states × 15 replicates × 1 million transmissions = 9 billion entries, about 1TB. [3] Size of summary data: 12 cells × 51 states × 15 replicates × 365 days × 90 health states × 3 counts = about 1 billion entries, 2.5GB.

Each of our workflows represent an interesting mix of data and compute intensive steps and thus crucially need HPC resources. Table I summarizes some of the key numbers for case studies we have described to illustrate the workflows. The partitioning of tasks and specific computation is carefully managed to reduce the amount of data transfer between the two clusters and achieve a near real-time response. Our paper advances the use of parallel and distributed computing in this important area – to the best of our knowledge this is the first time two HPC resources have been used in this manner to support near real-time epidemic planning and response.

**Table I.** Representative examples of individual workflows, their scale, and size of raw and summarized data generated by them.

| Workflow | # Cells | # States | # Replicates | # Simulations | Raw Output | Summ. Output |
| --- | --- | --- | --- | --- | --- | --- |
| Economic | 12 | 51 | 15 | 9180 | 3.0TB | 5.0GB |
| Prediction | 12 | 51 | 15 | 9180 | 1.0TB | 2.5GB |
| Calibration | 300 | 51 | 1 | 15300 | 5.0TB | 4.0GB |

## II. Description of the workflows

Figure 1 describes the overall workflow and how it is orchestrated across the two systems. Here we discuss three specific epidemiological workflows that are described in Figures 3, 4 and 5. Figure 2 shows the timeline of tasks involving human effort.

### Counter-factual analysis workflow (Figure 3)

Counter-factual analysis refers to the study of outcomes under various posted scenarios. The range of scenarios considered reflect the possible trajectory of the epidemic and is not known in advance. Our counter-factual analysis usually comprises various lockdown policies, compliances, and non-pharmaceutical interventions. The system is calibrated to reflect the current conditions on the ground. Usually such an analysis entails running a large factorial design and then computing certain outcomes that combine the output of the simulations and detailed synthetic social network, demographic and socio-economic data.

### Calibration workflow (Figure 4)

Calibration refers to finding plausible configuration(s) that produce simulation output similar to observed ground truth. Generally, such parameter searches are carried out by first defining a parameter space consisting of plausible parameter values, then evaluating the *closeness* of the simulation output to the ground truth at various points in that parameter space. However, when running the simulation is expensive, an emulator can be used in place of the actual simulation inside a calibration loop. An emulator is a statistical model that maps the input to the output of the simulation; it is cheap to run, and offers a way to quantify uncertainty for a deterministic system. To calibrate EpiHiper, a Gaussian Process [43], [46] emulator is used inside a Bayesian calibration framework for multivariate output [18], [29] to produce a set of plausible parameter configurations conditioned on the ground truth and associated uncertainty on the future predictions. The calibration task is carried out using the GPMSA framework [23] in Matlab. The calibration workflow typically resumes when ground truth data is updated or when we want to improve our predictions with a more appropriate parameter space or better-modeled mitigations. The calibration may reuse the existing model configurations, or generate new configurations as simulated by EpiHiper. This can vary by state. After simulation and aggregation, the time series of simulated case counts is compared to the ground truth with the aforementioned Bayesian approach to generate configurations for the prediction workflow.

### Prediction workflow (Figure 5)

To make predictions, we run simulations using the model configurations generated from the calibration workflow, and aggregate individual-level output to obtain future counts for various forecasting targets (e.g. confirmed cases, hospitalizations, deaths) at various spatial resolution (state or county level) with different temporal horizons (from one week to five months ahead) depending on the objective. The ensemble of the model configurations and the simulation output provides uncertainty quantification on the predictions. The prediction workflow typically resumes when the calibration workflow generates a set of model configurations, which are simulated by EpiHiper. The output is aggregated and analyzed by public health domain experts to identify inconsistencies (which may then trigger the calibration workflow again). If the predictions are deemed reasonable, we expand the configurations with a few possible future *what-if* scenarios (e.g. what if the stay-at-home order is lifted earlier; what if the mitigation compliance rate increases; what if testing and contact tracing are improved). Then simulations are run for the expanded configurations, and the results are combined with the as-is predictions.

## III. Description of Hardware, Software and Data

In this section, we describe the individual components of the overall workflow: (*i*) the underlying hardware, (*ii*) the software components used, and (*iii*) data used as input and generated as output. As mentioned earlier, the hardware consists of a home cluster and a remote super-computing cluster. Our typical workflow depicting the sequence of computations and data transfers between the two clusters are described in Figure 1, and described in more detail in the following section. The workflow relies on two important datasets as inputs: (*i*) the synthetic population and associated social contact network for the US, and (*ii*) COVID-19 specific disease parameters. Ranges for these parameters are based on best estimates from COVID-19 literature. The final component of the pipeline is the simulation-based models. Although we use meta-population models in addition to agent-based models, we will focus here on the agent-based models due to the significant computing challenges they pose.

### Home cluster and remote super-computing cluster

Our methodology makes use of two computing clusters, which we refer to as the *home cluster* and the *remote super-computing cluster*. The home cluster refers to the computing cluster available at the author’s home institution, Rivanna HPC Facility at University of Virginia. The home cluster is modest-sized relative to the significantly more powerful remote super-computing cluster, Bridges HPC Facility at Pittsburgh Supercomputing Center. The actual simulation runs are executed on the remote super-computing cluster. We find this distinction to be fairly typical and important, as most institutions do not have a super-computing facility available on their local premises, and researchers/practitioners often run the less compute-intensive parts of their workflows on their local systems, while running more computationally heavy tasks at dedicated super-computing facilities. Making this distinction explicit allows us to formally take into consideration issues arising from these kind of setups.

Note that commercial cloud computing platforms, such as Amazon EC2 and Google Cloud Platform, also provide services to make it relatively easy to set up computing clusters with software stacks mimicking those of HPC and super-computing facilities. Hence, this logical separation of “home cluster” and “remote super-computing cluster” is also relevant for institutions making use of hybrid cloud infrastructures, where a small local compute cluster is used alongside off-premises, cloud-based systems.

### Input Data: Synthetic populations and contact networks

Our epidemic computational models depend upon *detailed synthetic populations* and *contact networks* to support accurate and realistic simulations. Such data is prepared for each state, see Figure 6 for a summary of node and edge counts by US state.

**Figure 6.**
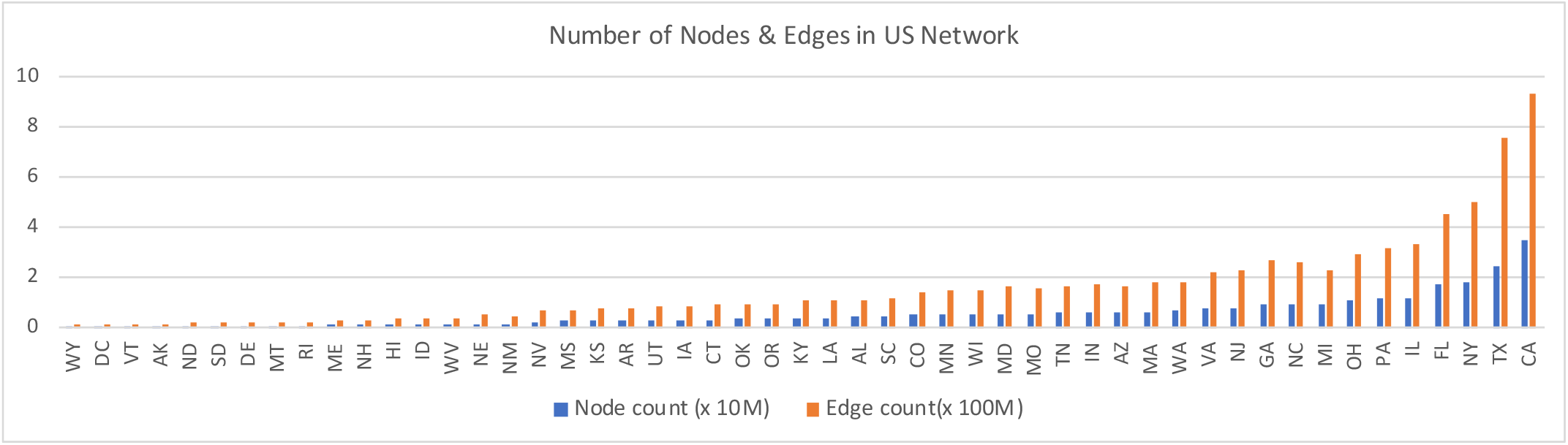
The diagram shows the number of nodes and edges in the contact network for each U.S. state as used in the simulations.

For each population, data is supplied as a comma-separated values (CSV) file containing the *traits* of each synthetic person. Whereas particular sets of traits may vary across simulations, typical choices for the US include household ID, age and age group, gender, county code, and the latitude and longitude of home locations. For design reasons, but also to avoid the cost of parsing and reading files from the file system during simulations, the population data is loaded into a PostgreSQL^4^ database server. All simulations access the population data by communicating with the database server at run-time.

The agent-based models use dynamic contact networks to encode interactions between persons during simulations. The initial dynamic contact network in EpiHiper is generated statically. However, during the course of the simulation, each edge in the contact network can be turned on and off dynamically as required in response to, for example, social distancing interventions. Like the person data, the contact network of each population is supplied to the simulations as one CSV file. Each edge in the contact network includes the identifiers of the two persons in contact, and is annotated by the start time and duration of the interaction, in addition to the context in which the persons meet (home, work, shopping, other, school, college, and religion). These contexts may not be the same for both persons, however; for example, if one person is at the store, their context may be shopping, while the grocer they came in contact with would be working. Due to the large size of the contact networks, the network is partitioned between different MPI processes at the beginning of the simulation run. The overall objective is to split the contact network such that each partition contains approximately the same number of edges, while, at the same time, ensuring that all incoming edges of any given node are in the same partition. In the current implementation, we utilize a simple algorithm to partition edges: given a partition, continue to allocate nodes to that partition until the number of incoming edges is greater than a threshold (*E/P* + *E*) where *E* is the number of edges, *P* is the number of partitions, and *E* is the tolerance factor. Note that even a simple partitioning scheme (such as the one described) takes a significant amount of compute time. This is why we use our current (simple) algorithm rather than one that is more sophisticated or optimal. We can also cache the result of the partitioning computation on disk, which saves time on future runs.

### Input Data to simulation: Disease progression parameters and parameter configurations

The disease model used for this work is shown in Figure 12 in the extended version of this paper (see footnote 2, page 1) and depicts the transmission of COVID-19 through interactions between individuals, and the subsequent disease progression of an infected individual. As shown in Figures 4 and 5, both calibration and prediction workflows start by generating simulation configurations, also known as *cells*. For calibration workflows, a larger number of cells are created, each with smaller numbers of replicates relative to routine prediction workflows, in order to explore the model configuration state space. For prediction workflows, however, a much smaller number of cells are generated which are based on the most likely model configurations from the calibration phase, each with a relatively larger number of replicates. The model configurations specify which populations and contact networks to use, as well as the disease parameters, interventions, initializations, and the number of days to simulate.

**Figure 12.**
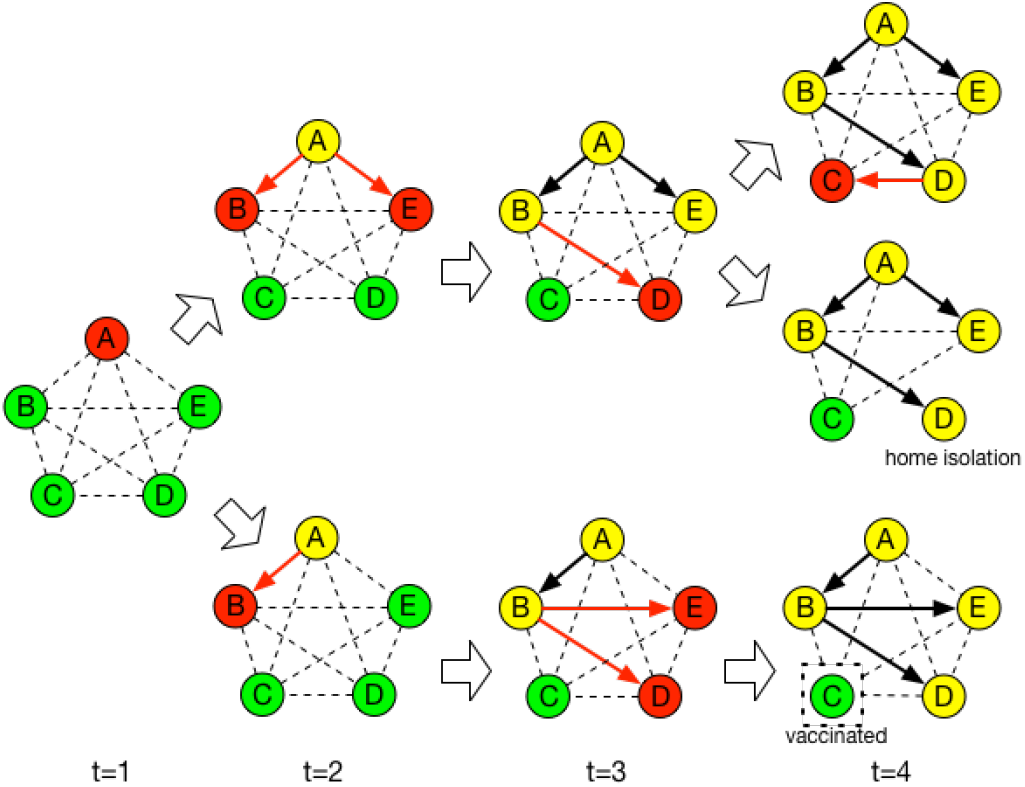
The COVID-19 Disease Model. This disease progression model is represented as a probabilistic timed transition system (PTTS): the state transitions are probabilistic, and, in many cases, are timed, i.e. transition after a given time period.

**Figure 13.**
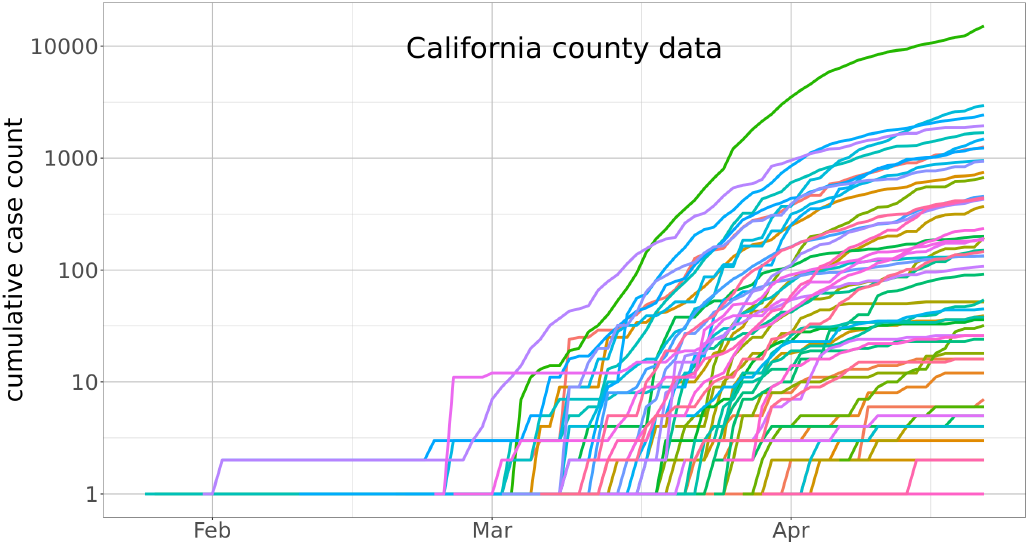
Time series of county-level cumulative confirmed cases of COVID-19 in California. Each state-level cumulative curve is obtained by summing its underlying county curves. There are a total of 3140 counties in the US.

**Figure 14.**
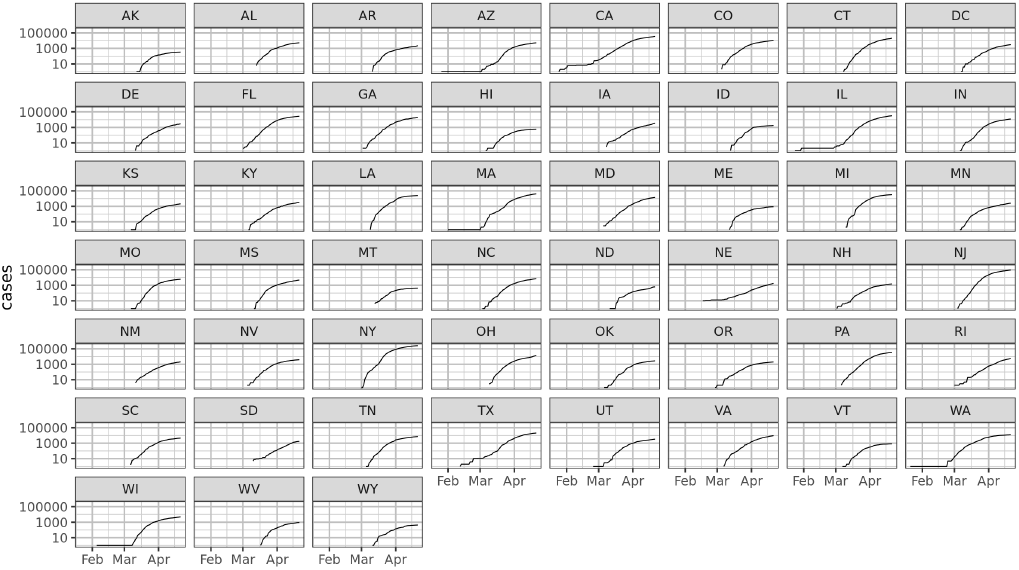
Time series of state-level cumulative confirmed cases of COVID-19. The incidence curves are highly noisy and often time-delayed. As a result, they pose a significant challenge when calibrating the simulations.

### Input data to calibration

For calibrations, we use confirmed cases from multiple data sources^5 6 7^ as our ground truth dataset. The ground truth data has county-level daily confirmed case counts starting from January 21, 2020, for over 3000 counties (as of April 22, 2020, there were 2772 counties with case counts greater than zero).

### Simulation-based models

*EpiHiper* is an agent-based discrete time simulation model for infectious disease spread in a social contact network. It is implemented as a parallel codeset in C++/MPI. It computes probabilistic disease transmission between nodes (representing individuals) in a network of edges (representing interactions between individuals), as well as the disease progression within each infected individual. It is based on the synthetic populations, accessible to the simulations via a database launched at run-time, and the synthetic contact network, partitioned pre-simulation and loaded into memory of the allocated processing units in order to support scalability. The simulation keeps track of the health state of each individual at each tick (the temporal resolution, set to one day in this case).

### Output data: dendograms and summary information

EpiHiper produces state transitions of all persons during the simulation. Each line of the output file written by EpiHiper includes the tick of the transition event, the identifier of the person, their exit state, and the identifier of the person causing the state transition in the case of disease transmission. The size of the output depends on the total number of ticks, overall epidemic size (number of infected persons), as well as the complexity of the finite state machine. Dendograms are part of this output, which are transmission trees rooted at initial infections.

From the individual-level output data, we can aggregate simulation results to the county level for different health states, and use the summary data for calibration and prediction. For example, the time series of daily cumulative counts of symptomatic cases at the state or county level are compared to the ground truth data as part of our calibration, and daily counts of symptomatic cases, hospitalizations, ventilations, and deaths are used in our predictions.

## IV. Orchestration of the workflows

### Structure of simulation jobs

The software stack on the remote super-computing cluster uses the Slurm scheduler for scheduling jobs, and Intel MPI for distributed communication. PostgreSQL servers are utilized to run the population databases. The number of processes to use per compute node is predetermined statically based on the configuration of individual compute nodes on the cluster. Furthermore, as described earlier, the population networks are partitioned statically beforehand, and they also determine the number of compute nodes/processes that will be utilized when running simulation jobs that use them. For simulations sharing a given user population, a single PostgreSQL server is started on a compute node and made available. The simulations use them to load population information at run-time. The data transfer between the home cluster and remote super-computing cluster utilizes the Globus platform^8^.

Every 24 hours, simulations are generated and executed to support the decision-making processes of policymakers. The process begins with the generation of the simulation configurations. The nature of the configurations generated depends on whether the calibration or prediction workflows are to be executed. Calibration workflows typically generate a large number of different model configurations to explore the space of the configurations. Prediction workflows, however, typically have a smaller set of model configurations, each replicated multiple times.

Once the configurations are generated, their transfer from the home cluster to the remote super-computing cluster is started manually using the Globus platform. Once the configurations are copied over, the population databases are started, one per population. To speed up the start of the population databases, snapshots of the databases are generated when the populations are initially created, and these snapshots are instantiated at run-time. Next, scripts are used to submit Slurm job arrays, which are scheduled to run using the heuristic scheduling strategy discussed above. Once simulation jobs have completed, the summary of simulation outputs are generated and transferred back to the home cluster using the Globus platform.

## V. Mapping and scheduling jobs on PSC Machines

Mapping our workflows on the remote supercomputing cluster is an important component. First, recall that the overall efficiency of the workflow is measured as time to complete the workflow rather than a single replicate of a single cell. Abstractly, our workflows can be thought of as large scale hierarchical *statistical experimental designs*. Each workflow is comprised of 51 regions (50 states and DC), and each region is then comprised of a number of cells that each denotes one combination of various parameters used to study a given problem. Each cell is further comprised of a number of replicates. Together, this represents a 3-level hierarchy: *regions-cells-replicates*. Each cell for a given region uses exactly the same input data; thus, we view our atomic jobs as ⟨*cell, region* ⟩. For certain workflows, it is more convenient and efficient from a scheduling perspective to group several cells into one to create jobs of appropriate sizes.

In general, the running time for a single replicate for ⟨*cell, region*⟩ is not fixed; this is due to (*i*) randomness within the computation, (*ii*) triggered interventions that can, at certain times, cause new calculations to be spawned based on the epidemic, (*iii*) number of processors assigned to the replicate and (*iv*) machine-specific randomness due to processors’ computation, access to the database etc. Nevertheless, by running the replicate several times we can obtain a reasonable bound on these times. For the workflows considered, we fixed the number of processors assigned to each ⟨*cell, region*⟩. We state the mapping problem in two stages:

### The workflow mapping problem (WMP)

We are given a set of ⟨*cell, region* ⟩ tasks, denoted by task *T* [*r, c*]. We assume that we know a bound *t*_*l*_(*T* [*c, r*]) and *t*_*r*_(*T* [*c, r*]) denoting the lower and upper bound on the time to complete task *T* [*c, r*] using *p*(*T* [*c, r*]) processing units. We use 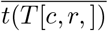 to denote the empirical mean running time obtained by running the computation several times and will use this for the rest of the paper. We assume *p*(*T* [*c, r*]) is known a priori. The problem is assigning an order to these tasks, then supplying this ordered set to the Slurm scheduler in such a way that minimizes the overall completion time of all tasks.

WMP is **NP**-hard. This can be seen by reducing the 2D Bin packing problem to the WMP problem: Rectangles become tasks: their width becomes *p*(*T*[*c,r*]) and their height is running time 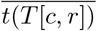. This reduction is useful and this correspondence also leads to natural heuristics for the problem discussed later in this section.

### Database Access Constraints

There is one additional constraint that needs to be taken care of which makes the problem computationally challenging. The constraint relates to database access. Recall that each task needs access to the input synthetic network. The number of simultaneous connections to the database are upper bounded for technology and efficiency reasons. We can capture this by using a compatibility graph. Usually compatibility constraints for tasks are captured as a coloring problem: we have a node for each task, and two tasks *u* and *v* have an edge iff they cannot be scheduled at the same time. A valid coloring captures a feasible schedule. In our case, the problem is more challenging and can be best described as a *new* kind of vertex coloring problem, which we will call a *relaxed coloring problem* (r-relaxed-coloring): We are given a graph *G*(*V, E*). Edges represent conflicts, and vertices represent tasks. We are given a number *r*. The (r-relaxed-coloring) is to assign a color to each node in the graph (such a graph would be constructed for each region separately) such that if a node *v* gets color *c*[*v*] then no more than *r* of its neighbors can get the color *c*[*v*]. If *r* = 1, we get the classical coloring problem and thus all the hardness results hold for the relaxed coloring problem as well.

### The DB-access constrained workflow mapping problem (DB-WMP)

DB-WMP is a constrained version of the WMP wherein the number of tasks that can be scheduled simultaneously is bounded. Thus the general DB-WMP problem can be thought of as 2D Bin packing with an interesting compatibility constraint.

### Our Mapping heuristic (MAP)

Our mapping heuristic is based on a few simplifying assumptions and exploiting the problem structure. *Assumption 1:* We assume that all tasks for a given region take the same amount of time which is 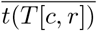, in other words 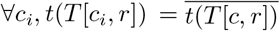. *Assumption 2:* All tasks have to be scheduled non-preemptively. *Assumption 3:* The number of connections that can be made by tasks corresponding to a region *r* is bounded by *B*(*T* [*r*]) (i.e. it is not dependent on the cell). *Assumption 4:* For each region, all the tasks *T* [*c*_*i*_, *r*] require the same number of threads for simplicity and are denoted by *dt*(*T* [*c, r*]), thus Σ_*c*_ *dt*(*T* [*c, r*]) *> B*(*T* [*r*]). Our heuristic is motivated by the non-decreasing first fit heuristic. Recall the task of this heuristic is to provide the Slurm scheduler an ordering and chunking of tasks. Slurm further does a certain amount of real-time optimization. It comprises of the following steps:

**Step 1** Split the overall database so that we have one database per region. For various system-level reasons and from the standpoint of human productivity, each such database occupies one node of the system. Thus, all tasks corresponding to a given region can access the region-specific database. Access by each region can now be done in parallel with no constraints beyond the fact that we have a constraint on the total number of processors. Let *T* [*r*] = *∪* _*c*_*T* [*c, r*] denote the set of tasks for region *r*. The above decomposition makes the coloring problem easy. We now have *r* subsets — one subset per region. There is no edge between the subset, and the graph within each subset is a complete graph. All tasks for a given region *r* thus belong to a Region set *RS*(*r*).

**Step 2**. Organize the tasks in non-increasing order by time needed to complete the computation. The time is directly correlated with the size of the network for each cell. Using an idea motivated by the 2D Bin packing methods, we use a level-oriented approach [11], [12], [47], [55]. Think of processors on the X-axis and time on the Y-axis. The tasks are mapped from left to right (in terms of available processors), in rows forming levels. Within the same level, all tasks are packed so that their bottoms align. The first level is the bottom of the strip and subsequent levels are defined by the time taken of the slowest task on the previous level.

**Step 3**. We considered two different mapping algorithms: The Next-Fit Decreasing time with database constraints (NFDT-DC) algorithm assigns the next task *T* [*c, r*] (in non-increasing time) on the current level if *T* [*c, r*] fits and database access constraints are satisfied. Otherwise, the current level is “closed” and a new level is created. The First fit decreasing time with database access constraint (FFDT-DC) algorithm schedules the next task in non-increasing order of time, until either the database access constraint for the region is violated, or, if no level can accommodate the task, a new level is started.

Without the database constraints, the NFDT-DC and FFDT-DC algorithms have worst-case performance guarantees of 2 and 17*/*10 respectively. Let *EC* denote the empirical efficiency of our method. This is computed as the ratio of the total time used by all processors as they were computing divided by the product of the total processors and the time when the last task was completed. As the next section discusses, our algorithms do quite well; the FFDT-DC ordering achieves a very high system utilization.

## VI. Performance Analysis

### Runtime performance of EpiHiper

Figure 7 (top) shows that EpiHiper’s running time increases linearly with its input size. On the other hand, Figure 7 (middle) shows how increasing the number of processing units for three medium-to-large networks can significantly improve simulation performance. The improvement in the performance, however, starts to decrease beyond a certain number of processing units due to increasing communication costs between processes. It may even become slower with too many processes. In Figure 7 (bottom), we show that EpiHiper’s running time depends also on the interventions implemented in the simulation. In the base case, the simulation has implemented VHI (voluntary home isolation), SC (school closure), and SH (stay-at-home). When we add more interventions to the simulation, the running time increases. The simpler interventions RO (partial reopening), which extends SH, and TA (testing and isolating asymptomatic cases), which extends VHI, increase running time marginally. The more complex interventions PS (pulsing shutdown), which repeatedly alternates SH and RO, and D1CT (distance-1 contact tracing and isolating), which affects many more nodes and edges, significantly increase the running time. The most complex intervention we have implemented so far, D2CT (distance-2 contact tracing and isolating), increases the running time by almost 300% from the base case.

**Figure 7.**
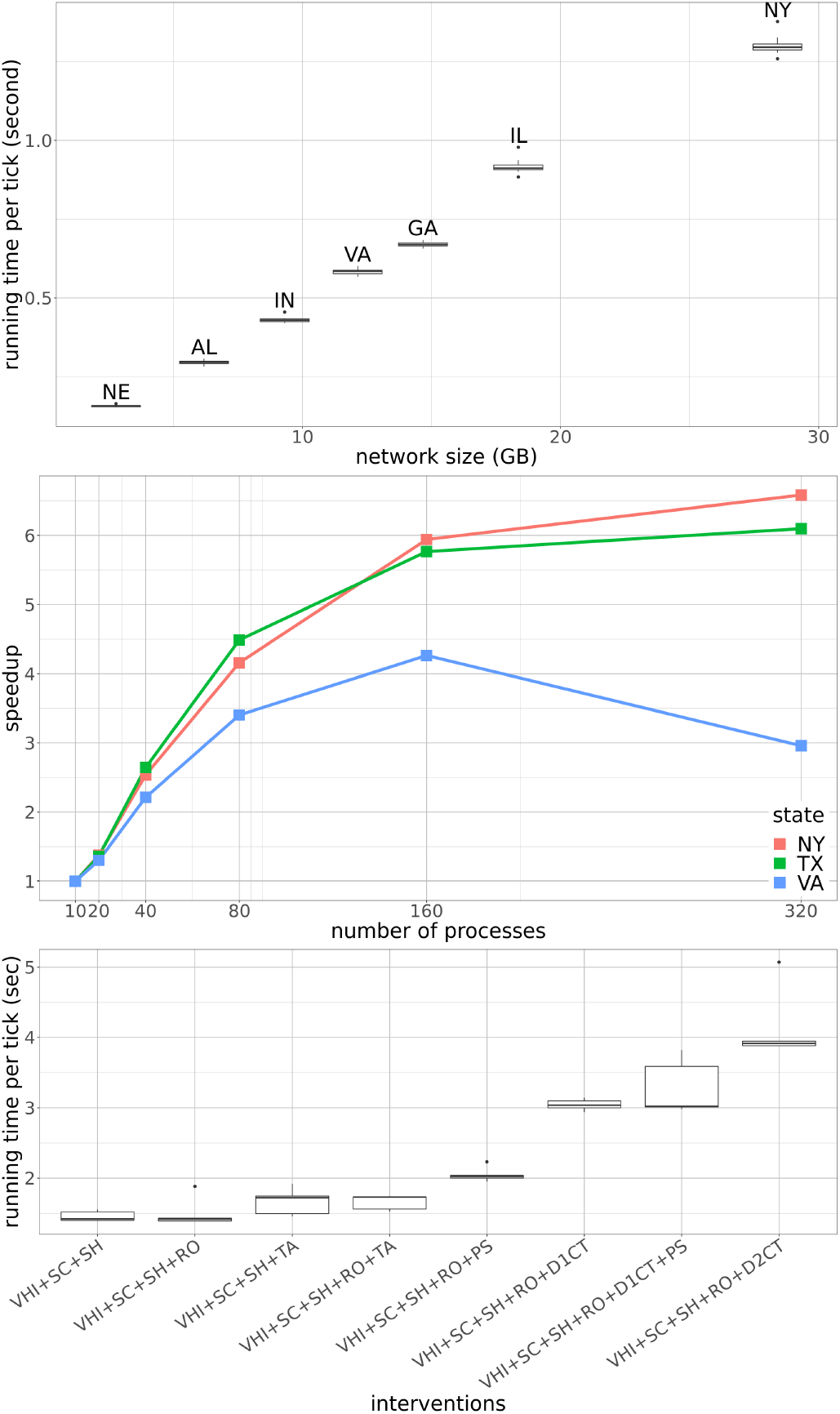
(top) Running time of EpiHiper on networks of different sizes given 40 processing units. (middle) As the number of processing units increase, the corresponding improvement in the performance of the simulations illustrates the strong scaling results of EpiHiper. Beyond some point, which varies with the problem size, the benefit of using more resources starts to diminish. (bottom) Running time of EpiHiper varies with different interventions in the simulation. Simulations with more interventions, or with more complex interventions, take more time.

### Scheduling and partitioning simulation jobs

The primary purpose of the workflow presented in this paper is to serve the needs of policymakers by providing them with timely predictions of disease progression that incorporates the most recent data. To serve this purpose, we face a high throughput problem where we have to maximize the number of simulation jobs we can execute in order to generate calibration and projection results. We are given two constraints (*i*) limited compute time (10:00pm −8:00am), and (*ii*) limited number of compute nodes as described in (Table II). The job scheduling strategy presented in the previous section focuses on timeliness, that is, reducing the time span required to execute a given set of jobs on the compute cluster.

The minimal memory requirement per job is given by the size of the contact network which is stored in memory during runtime. Furthermore, the memory requirements may increase due to the complexity of interventions performed in a scenario. Our experience is that in nearly all cases, the additional memory is proportional to the network size. For simplicity, we therefore divided the 51 regions (networks) into 3 categories: small (2 compute nodes), medium (4), and large (6). With these assignments, we were able to guarantee that the jobs have sufficient memory to complete even the complex intervention scenarios. We intentionally avoided using partial nodes in order to limit problems caused by competing memory requirements of different jobs running on the same node. By consistently using the maximum number of cores available per node, we ensure that the available compute resources are fully utilized.

Furthermore, we chose to create static network partitions in order to save compute time, since partitioning the network to binary chunks for California alone would take over one hour. The time for partitioning a network is larger than the typical run time for a simulation run, which usually requires between 100 to 300 time steps of about 3 seconds each for a network the size of California (Figure 7 (left)). We chose not to assign additional resources, since Figure 7 (right) shows that increasing the number of compute nodes for a single simulation gives diminishing returns in terms of runtime; due to that, the cost of messaging negates any gains obtained from using more compute power. After the general categorization of all jobs into the 3 categories above, we are faced with maximizing the number of simulations we are able to run within our time window. Every night, we have a varying number *N* jobs to run, and face the challenge of scheduling them efficiently.

While supercomputing facilities can grant access to a large amount of resources, access to these resources come at a large cost, either for the users who are paying for the resources directly or for the taxpayer in case of publicly funded research. Thus one important metric to consider for the scheduling problem is the issue of resource utilization. Figure 9 shows the resource utilizations for our workflows, in terms of percent of CPU hours allocated that were actually used. The Figure 9 (left) shows the distribution of utilizations of 9 workflow runs which simulated all 51 regions, while Figure 9 (right) shows the same for 24 workflow runs that simulated multiple configurations for the state of Virginia. In the case of all state workflows, we have a median utilization of 96.698% while the same for Virginia-only workflows is 95.534%. Note that the above results are for the scheduling configuration (FFDT-DC) where the largest jobs were scheduled first. Our initial workflow runs without this scheduling scheme (NFDT-DC) led to utilization numbers between 44.237% and 55.579% for the all-state case.

**Figure 9.**
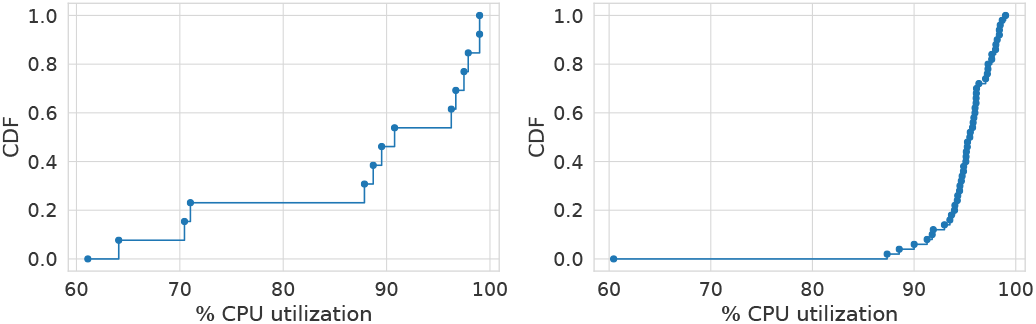
Utilization of compute resources on remote super computing clusters for different days of workflows. The *left* figure shows utilization for the days when all 50 states and DC were simulated, while the *right* figure shows utilization for days when only different cells for the state of Virginia were simulated.

**Table II.**
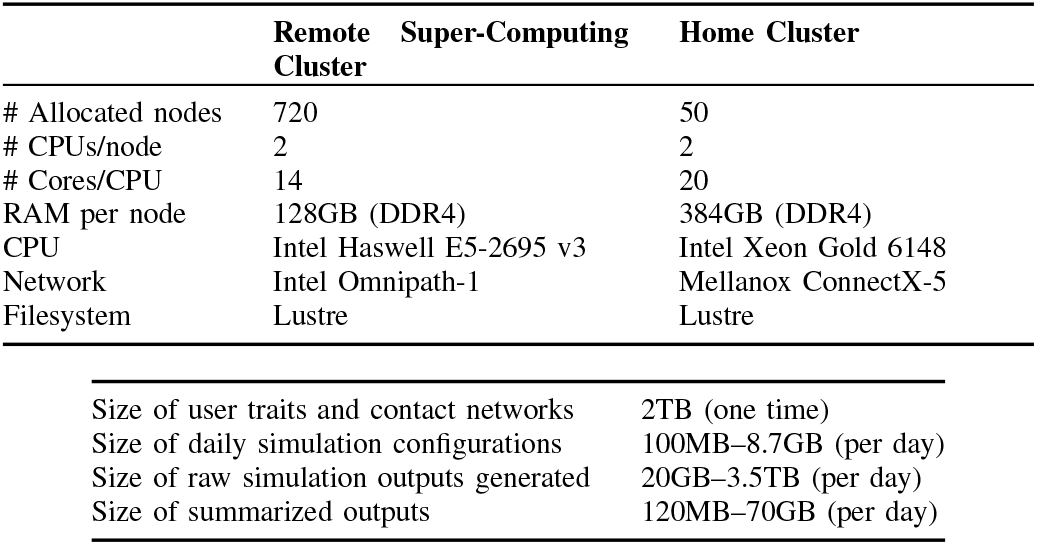
Configuration of the remote super-computing cluster (Bridges HPC Facility at Pittsburgh Supercomputing Center) and home cluster (Rivanna HPC Facility at University of Virginia), along with data generated and moved across them.

### Runtime performance of simulation ensembles on remote system

Here we present the runtime characteristics of the simulations. Figure 8 shows the variance in runtime (across compute nodes) for the 50 US states and DC for a single representative day of simulation. To understand the dynamic nature of the total memory required for the different EpiHiper simulations, we plot the total memory required for different cells, and US states in Figure 10. Figure 8 shows that runtimes of simulations are dependent on intervention scenarios and is strongly correlated to the network size. The memory increase during the simulation in Figure 10 is due to the intervention scheduled at fixed time points. Figure 10 (left) shows that memory requirements in the same scenario may depend on the compliance of nodes with the interventions, i.e., higher compliance and, therefore, more scheduled changes to the system state require more memory. Finally, Figure 10 (right) shows that the final memory requirements are strongly correlated with the initial requirements, i.e., the network size.

**Figure 8.**
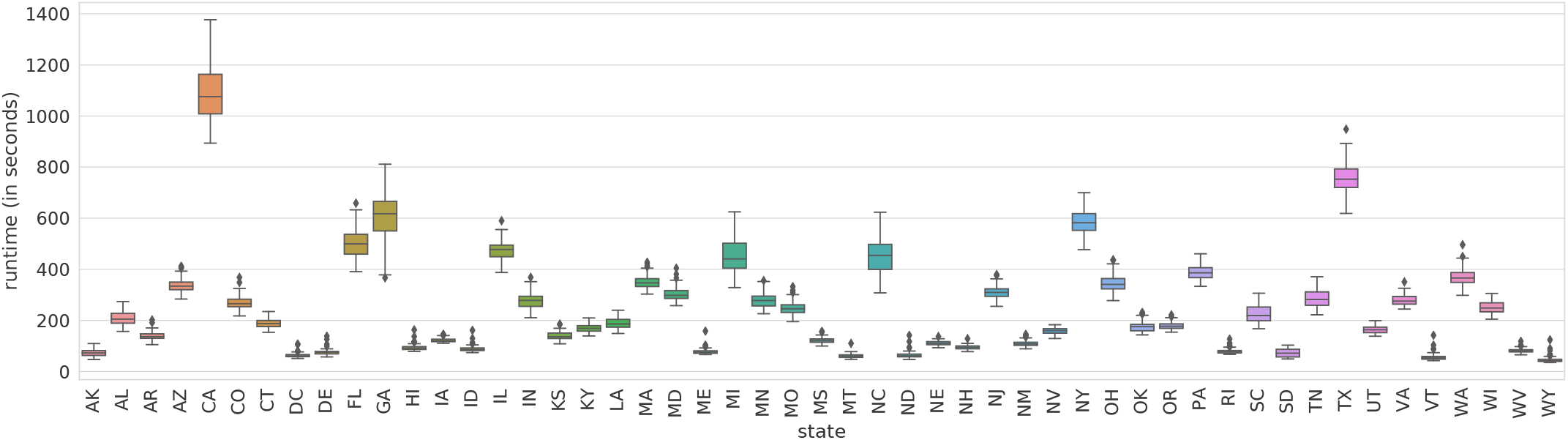
Variance in runtime for EpiHiper simulations for different US states, across different cells or simulation configurations.

**Figure 10.**
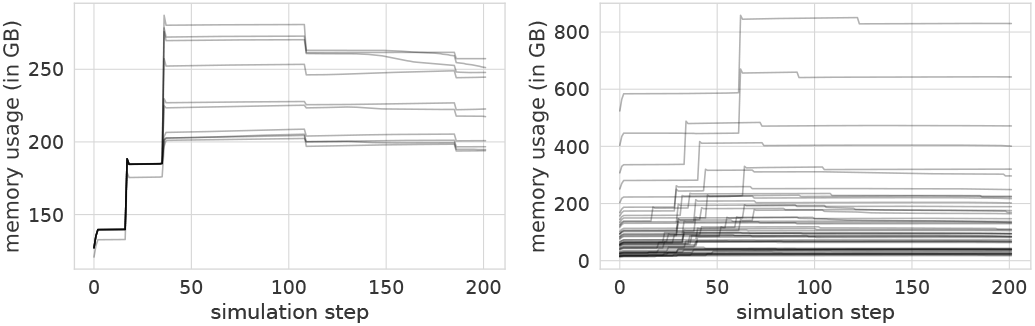
Changes in memory required for different cells for VA (left) and US states (right) at different timesteps. In the left figure, every line corresponds to a different cell or simulation configuration generated for the state of Virginia, and shows the mean memory required (across replicates). In the right figure, every line corresponds to a different US state, and shows shows the mean memory required (across cells).

## VII. Illustrative case studies

In this section, we discuss three case studies: (*i*) medical cost of the pandemic illustrated in Figure 3, (*ii*) forecasting workflow illustrated in Figure 5 and (*iii*) calibration workflow illustrated in Figure 4. The second and third workflows are discussed in the Appendix F.

The workflows selected illustrate a range of tasks undertaken using the two supercomputing clusters.

### Case study: Medical costs of COVID-19

In this study, we estimate the medical costs of COVID-19 in the US. The overall impact also includes the cascading effect to the Gross Domestic Product (GDP), which can be analyzed by an input-output or general equilibrium model. Since the purpose is to demonstrate the workflow, we will focus on the medical cost estimating.

The medical costs include costs incurred by COVID-19 patients for medical attention, hospitalization, ventilator support, etc. For each patient, the total costs depend on the disease severity. We consider a calibrated (towards *R*_0_ = 2.5) disease model with different scenarios with respect to NPI (non-pharmaceutical intervention) duration and compliance. For each scenario in our factorial design of 12 cells, we run simulations with 15 replicates for each of the 51 regions (50 states and DC), with county-level seeding derived from county-level confirmed case counts. The simulation outputs individual-level data on who are infected, receiving medical attention, hospitalized, and/or on ventilator support each day. The aggregate data is used to compute the total medical costs for each scenario. The details of the study are described in [9]. The workflow for our economic impact analysis consists of the following steps: *(i)* On the local or remote cluster, calibrate the disease model towards *R*_0_ = 2.5. *(ii)* On the local cluster, prepare simulation configuration files for a factorial design of different NPI durations and compliance; get the most recent county-level confirmed case counts and use them to prepare county-level seeding. *(iii)* Send the disease model, seeding, and configuration files to the remote cluster. *(iv)* On the remote cluster, create database jobs and simulation jobs, use our scheduling heuristic to submit jobs, and run post-simulation data aggregation. *(v)* Transfer aggregate simulation data to the local cluster. On the local cluster, run the economic impact model to estimate medical costs.

## VIII. Related Work

Over the last decade, there has been substantial interest in developing scalable solutions to support various epidemiological tasks. This includes: planning and counterfactual analysis, forecasting, and various resource optimization problems. There has also been interest in developing web-based tools to support these tasks. The models used in these papers often range from simple statistical models to compartmental models. Due to space considerations, we only highlight a few important papers here.

Agent-based models in epidemic sciences can be traced back to the earlier work on human immunodeficiency virus (HIV), although the models were largely focused on the structural analysis of small networks; see [16], [21], [24]. The use of the models was largely restricted to modeling studies. Recent papers that aim to scale these simulations to the national level include [5], [42].

In [3], [14] the authors report on the development of web-based systems to carry out large computational experiments in support of epidemic planning. See [15], [26], [40] for other related efforts.

Researchers have also created data-driven pipelines to support epidemic forecasting. CDC runs an annual challenge in this area for studying influenza. Several important advances have been made to improve the overall forecasts; most of the work in this space is either statistical time series models or simple compartmental mass action models; see [44], [48]. Operational agent-based models for epidemic forecasting have not yet been reported on. Recently there have also been a lot of community-wide efforts related to COVID-19^9 10 11^; our group submits forecasts to a number of these efforts.

Developing scalable pipelines and workflows for HPC tasks involving large datasets has also been well-studied in literature [19], [27], [33], [41]. For example, the authors of [19] present a technique for building scalable workflows for analyzing large volumes of satellite imagery data, while [33] present a system for analyzing workflows related to weather-sensing data. Other studies have presented generalized methodologies for building scalable workflows for tasks requiring HPC platforms [7], [27].

Recently there has been a flurry of papers on developing agent-based and equation-based models for planning and response to the COVID-19 pandemic; see [1], [2], [10], [20], [22], [30], [31], [36], [45], [54] The present paper does not focus on our agent-based models — they are covered in a companion paper.

Our primary focus is on creating scalable HPC-oriented workflows to support a range of epidemiologically relevant tasks in real-time. Our work shows how two large supercomputing clusters have been used to meet that goal, and is a step towards demonstrating the use of hybrid supercomputing cloud technology for epidemic science. The resulting challenges are unique, and form an important data-driven simulation platform.

## IX. Conclusion

We describe how we have developed high performance computing-oriented epidemic workflows in order to support the planning and response to pandemics such as COVID-19. Our workflows are unique in their use of two geographically separated supercomputing clusters. The workflows are also unique from the standpoint of executing large data-intensive steps that incorporate daily county-level surveillance and policy data, national and highly resolved agent-based simulations of epidemic processes, and post-simulation analytics for projections and counter-factual analysis. The work arose in response to requests from federal and state agencies to support their work on COVID-19 planning, and, using this approach, we have been able to provide uninterrupted support for over 30 weeks. This was accomplished in record time – we began this effort in early March after access to such machines was made possible by the HPC Consortium. We were provided with unprecedented support by Bridges HPC Facility at Pittsburgh Supercomputing Center. Our results demonstrate that *real-time data-driven high resolution epidemics science at national scale is possible*. COVID-19 is not over; we are witnessing a second, or possibly third, wave. The tools we have developed will assist policymakers in developing and evaluating new intervention measures, and will hopefully help prevent COVID-19 from becoming an even larger-scale outbreak.

## Data Availability

The data can be shared upon request

## Acknowledgment

We thank the anonymous reviewers for their valuable suggestions.

This work was partially supported by National Institutes of Health (NIH) Grant 1R01GM109718, NSF BIG DATA Grant IIS-1633028, NSF Grant No.: OAC-1916805, NSF Expeditions in Computing Grant CCF-1918656, CCF-1917819, NSF RAPID CNS-2028004, NSF RAPID OAC-2027541, US Centers for Disease Control and Prevention 75D30119C05935, University of Virginia Strategic Investment Fund award number SIF160, Google Grant, and Defense Threat Reduction Agency (DTRA) under Contract No. HDTRA1-19-D-0007. Any opinions, findings, and conclusions or recommendations expressed in this material are those of the author(s) and do not necessarily reflect the views of the funding agencies.

The authors would like to thank members of the Bio-complexity Institute and Initiative for useful discussion and suggestions. We thank the staff members at the University of Virginia’s high performance computing center for their help in ensuring our jobs run smoothly every day on the Rivanna Cluster. We also thank staff members at the Pittsburgh Supercomputing center (PSC), Dr. John Towns head of NSF XSEDE project and Dr. Shawn Brown, head of PSC for providing us the needed HPC resources at such a short notice; our work would not have been possible without their support. The timely and important resource provided by the NSF during this national crisis was crucial and is sincerely appreciated.

## Appendix A

### Networked Epidemiology

In this paper, we will focus on networked models, which consider epidemic spread on an undirected social interaction network *G*(*V, E*) over a population *V*, where each edge *e* = (*u, v*) *∈ E* implies that individuals (also referred to as nodes) *u, v* ∈ *V* interact. The specific form of interaction depends on the disease being modeled; e.g. sexually transmitted diseases require physical sexual contact, while influenza-like illnesses require physical proximity. Let *N* (*v*) denote the set of neighbors of *v*. In its simplest form, the SIR model on the graph *G* is a dynamical process in which each node is in one of *S* (susceptible), *I* (infected) or *R* (recovered) disease states. Infection can potentially spread from u to v along edge *e* = (*u, v*) with a probability of *β*(*e, t*) at time instant *t* after u becomes infected, conditional on node v remaining uninfected until time *t* — this is a discrete version of the rate of infection for the ordinary differential equation (ODE) model discussed earlier. We let *I*(*t*) denote the set of nodes that become infected at time t. The (random) subset of edges on which the infections spread represents a disease outcome, and is referred to as a dendogram. This dynamical system starts with a configuration in which there are one or more nodes in state I and reaches a fixed point in which all nodes are in states *S* or *R*. Some of the key challenges are: characterizing the evolution of |*I*(*t*) | with time (i.e., the epicurve) as a function of the disease model parameters and network structure; predicting the size of the peak, i.e., max_*t*_ |*I*(*t*) | and its timing; and how to reduce the outbreak size Σ_*t*_ |*I*(*t*) | by identifying effective interventions, such as vaccinating nodes (which can be modeled as node deletions), and social distancing between nodes (which can be modeled as edge deletions).

In Figure 11, we illustrate random disease propagation in a network and effects of interventions with three trajectories of disease evolution in a small contact network. The small network represents daily contacts between five people in a workplace or a school classroom. They are colored for their health states: susceptible-green, infectious-red, recovered-yellow. Infections start from A, which in one scenario infects B and E, in another scenario infects B only. In the former scenario, B infects D; then D either chooses to stay and infects C (so all five people have been infected in this trajectory), or chooses to go home for isolation (so avoids transmitting the disease to C). In the latter scenario, B infects D and E; while C decides to get vaccinated and avoids being infected.

**Figure 11.**
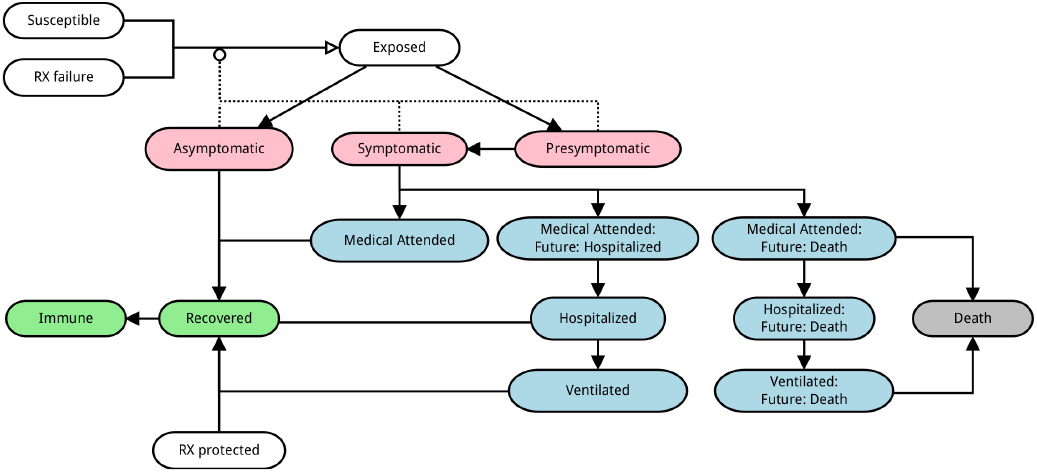
An example showing the stochastic disease propagation in a social network and the effect of interventions.

## Appendix B

### The Disease Model Parameters

The within-host disease transmission model is shown in Figure 12. **Transmission** occurs when an individual in one of the states *Susceptible* or *RX Failure* comes in contact with an individual in the states *Presymptomatic, Symptomatic*, or *Asymptomatic*. The individual transmissions are governed by the parameters in Table IV. **Progression** from one disease state to the next is governed by the parameters in Table III

**Table III.**
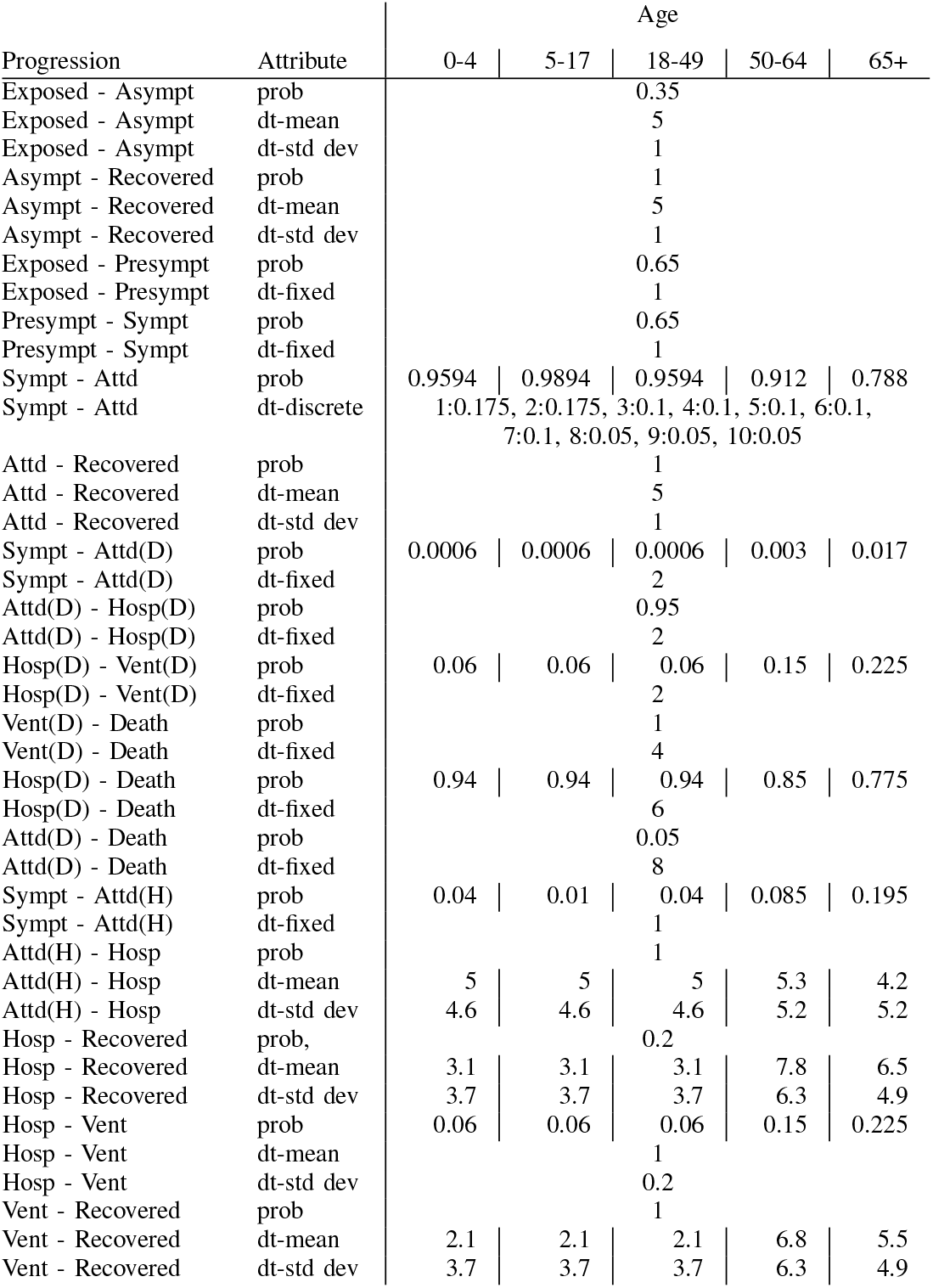
Disease progression parameters as given by the CDC document [8]. One value per line applies to all age groups. Abbreviations: prob: probability, dt: dwell time, Attd: attended, Hosp: hospitalized, Vent: ventilated, (D): resulting in death, (H): resulting in hospitalization

| Progression | Attribute | Age |  |  |  |  |
| --- | --- | --- | --- | --- | --- | --- |
|  |  | 0-4 | 5-17 | 18-49 | 50-64 | 65+ |
| Exposed - Asympt | prob |  |  | 0.35 |  |  |
| Exposed - Asympt | dt-mean |  |  | 5 |  |  |
| Exposed - Asympt | dt-std dev |  |  | 1 |  |  |
| Asympt - Recovered | prob |  |  | 1 |  |  |
| Asympt - Recovered | dt-mean |  |  | 5 |  |  |
| Asympt - Recovered | dt-std dev |  |  | 1 |  |  |
| Exposed - Presympt | prob |  |  | 0.65 |  |  |
| Exposed - Presympt | dt-fixed |  |  | 1 |  |  |
| Presympt - Sympt | prob |  |  | 0.65 |  |  |
| Presympt - Sympt | dt-fixed |  |  | 1 |  |  |
| Sympt - Attd | prob | 0.9594 | 0.9894 | 0.9594 | 0.912 | 0.788 |
| Sympt - Attd | dt-discrete | 1:0.175, 2:0.175, 3:0.1, 4:0.1, 5:0.1, 6:0.1, 7:0.1, 8:0.05, 9:0.05, 10:0.05 |  |  |  |  |
| Attd - Recovered | prob |  |  | 1 |  |  |
| Attd - Recovered | dt-mean |  |  | 5 |  |  |
| Attd - Recovered | dt-std dev |  |  | 1 |  |  |
| Sympt - Attd(D) | prob | 0.0006 | 0.0006 | 0.0006 | 0.003 | 0.017 |
| Sympt - Attd(D) | dt-fixed |  |  | 2 |  |  |
| Attd(D) - Hosp(D) | prob |  |  | 0.95 |  |  |
| Attd(D) - Hosp(D) | dt-fixed |  |  | 2 |  |  |
| Hosp(D) - Vent(D) | prob | 0.06 | 0.06 | 0.06 | 0.15 | 0.225 |
| Hosp(D) - Vent(D) | dt-fixed |  |  | 2 |  |  |
| Vent(D) - Death | prob |  |  | 1 |  |  |
| Vent(D) - Death | dt-fixed |  |  | 4 |  |  |
| Hosp(D) - Death | prob | 0.94 | 0.94 | 0.94 | 0.85 | 0.775 |
| Hosp(D) - Death | dt-fixed |  |  | 6 |  |  |
| Attd(D) - Death | prob |  |  | 0.05 |  |  |
| Attd(D) - Death | dt-fixed |  |  | 8 |  |  |
| Sympt - Attd(H) | prob | 0.04 | 0.01 | 0.04 | 0.085 | 0.195 |
| Sympt - Attd(H) | dt-fixed |  |  | 1 |  |  |
| Attd(H) - Hosp | prob |  |  | 1 |  |  |
| Attd(H) - Hosp | dt-mean | 5 | 5 | 5 | 5.3 | 4.2 |
| Attd(H) - Hosp | dt-std dev | 4.6 | 4.6 | 4.6 | 5.2 | 5.2 |
| Hosp - Recovered | prob, |  |  | 0.2 |  |  |
| Hosp - Recovered | dt-mean | 3.1 | 3.1 | 3.1 | 7.8 | 6.5 |
| Hosp - Recovered | dt-std dev | 3.7 | 3.7 | 3.7 | 6.3 | 4.9 |
| Hosp - Vent | prob | 0.06 | 0.06 | 0.06 | 0.15 | 0.225 |
| Hosp - Vent | dt-mean |  |  | 1 |  |  |
| Hosp - Vent | dt-std dev |  |  | 0.2 |  |  |
| Vent - Recovered | prob |  |  | 1 |  |  |
| Vent - Recovered | dt-mean | 2.1 | 2.1 | 2.1 | 6.8 | 5.5 |
| Vent - Recovered | dt-std dev | 3.7 | 3.7 | 3.7 | 6.3 | 4.9 |

**Table IV.**
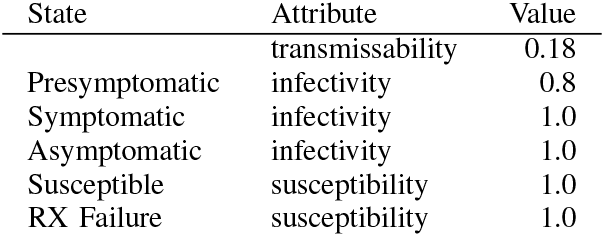
Disease Transmission Parameters

## Appendix C.

### Generating synthetic populations and networks

A synthetic population of a region may be regarded as a digital twin of the real population of the region. Here we provide a compact summary of the model and methodology behind constructing synthetic populations and their contact networks in the case of the US, see [38] for details. These populations and networks are central to the EpiHiper simulation model.

To construct a population for a *geographic region R* (e.g., Virginia), we first choose a collection of *person attributes* from a set (e.g., age, gender, and employment status) and a set 𝒯 _*A*_ of *activity types* 𝒟 (e.g., Home, Work, Shopping, Other, School). The precise choices of 𝒟 and 𝒯 _*A*_ are guided by the particular scenarios or analyses the population will serve. Described at a high level, we (*i*) construct people and places, (*ii*) assign activity sequences to people, (*iii*) map each activity for each person to a location (including the time of the visit), and (*iv*) from this, we derive a contact network using co-occupancy to infer edges. The construction is broken down in a sequence of steps outlined in the following.

Using *iterative proportional fitting* (IPF) [4], [13] the **base population** model constructs a set of individuals 𝒫 where each person has assigned demographic attributes from 𝒟. By design, this ensures that 𝒫 matches the actual distributions and Public Use Microdata Sample (PUMS) data from the U.S. Census [51], which is the input data for the model. Additionally, this model partitions 𝒫 into a set of *ℋ households*, where the notion of household encompasses the traditional notion of “family”, but also any other subset of individuals residing in the same *dwelling unit* (e.g., dormitories, army barracks, or prisons).

After household assignment, each individual *p ∈* 𝒫 is assigned a week-long activity sequence *α*(*p*) = (*a*_*i,p*_)_*i*_ where each *activity a*_*i,p*_ has a *start time*, a *duration*, and an *activity type* from 𝒜. Data sources used for this step include National Household Travel Survey (NHTS) [53], American Time Use Survey (ATUS) [52] and Medical Treatment Utilization Schedule (MTUS) [49]; these sources are fused to form consistent, week-long activity sequences. We write *α* : 𝒫 *→* 𝒜 for the mapping assigned to each person. For this construction we use Fitted Values Matching (FVM) for adults [32], and CART (Classification And Regression Tree) for children (see, e.g., [6]).

The **location model** constructs a set of spatially embedded locations ℒ consisting of *residence locations* where households live, and activity locations where people conduct their non-Home activities. This construction is highly granular and is rooted in data such as the MS Building data [37], HERE/NAVTEQ data [28] for points-of-interest (POIs) and land-use classifications, National Center for Education Statistics (NCES) [39] data for public schools, as well as LandScan^12^, OpenStreetMap^13^, and Gridded Populations of the World (GPW) v4^14^.

For each person *p ∈* 𝒫 the **location assignment model** assigns a location 𝓁= 𝓁 (*a*_*i*_) to each of their activities *a*_*i*_. We denote the sequence of locations visited by *p* as *λ*_*p*_ = (*λ*_*i*_)_*p*_. The location assignment model uses American Community Survey (ACS) commute flow data [50] to assign the target county *c* for Work activities, and a particular location randomly within *c* work weights assigned to each location in *c*. School activity locations are assigned based on NCES data, with remaining activities anchored near home and work locations.

Finally, the **contact network model** uses the location assignment to derive the bipartite *people location graph G*_*P L*_ with vertex sets *V*_1_ = 𝒫 and *V*_2_ = ℒ and a labeled edge (*p, 𝓁*) whenever *p* visits *𝓁* with labels activity type and time of visit. From this we derive the list of visitors to each location and the graph *G*_max_ with vertex set 𝒫 and edges all *e* = (*p, p*) for people *p* and *p′s* that are simultaneously present at the same location. Merely being present at a location at the same time does not imply a contact, and sub-location contact modeling is applied at each location to determine which of the edges of *G*_max_ should be retained to form the *contact network G*. Finally, for the applications and scenarios of this paper, we project from *G*, the week-long contact network, to *G*_Wednesday_, representing the contact network on a “typical day”.

## Appendix D

### The Agent-based Simulation Model

EpiHiper supports disease models which are comprised of *disease states, disease transmissions* (through contacts), and *disease progressions*. These disease models are specified independently from the people and their contact network among whom the diseases spread. In other words, all individuals have the same dynamics; that is, they have the same infection process and disease progression dynamics. All inputs to EpiHiper are given in JSON format, with the exception of the contact network, which, due to its large size, is in csv or binary format.

**Disease Transmissions** are caused by contact between a susceptible individual *P*^*s*^ in state *X*_*i*_ and an infectious individual *P*^*i*^ in state *X*_*k*_. The susceptible individual *P*^*s*^ will transition to an exposed state *X*_*j*_ based on information given by the transmission *T*_*i,j,k*_, the contact *E*(*P*^*i*^, *P*^*s*^), and attributes from the individuals *P*^*s*^ and *P*^*i*^ such as the susceptibility *σ*(*P*^*s*^) and infectivity *ι*(*P* ^*i*^).

Under the assumption of independence of transitions across contacts for individual *P*^*s*^ with infectious individual *P*^*i*^, the *propensity* of the state transition to the exposed state *X*_*j*_ based on the transmission *T*_*i,j,k*_ and the contact *E*(*P*^*i*^, *P*^*s*^) is defined as:

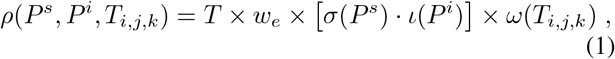

where *T* is the contact duration, *w*_*e*_ is the edge weight, and *ω*(*T*_*i,j,k*_) is the transmission rate.

The *propensities* of all state transitions to the exposed state *X*_*j*_ are summed. We use the Gillespie algorithm [25] to determine whether a transition occurs during the simulation interval, and which contact caused it.

**Disease Progression** covers the health state transitions within an individual *P* that are independent of other people. For the EpiHiper model, a disease progression diagram describes all the possible health state transitions that take place within a person. The nodes of the diagram are the health states *𝒳* = {*X*_*i*_} and directed edges of the form *e* = (*X*_*i*_, *X*_*j*_) with an assigned probability *p*_*e*_ = prob(*X*_*i*_, *X*_*j*_) and a *dwell time distribution D*_*e*_. The sum of all probabilities of transitioning out of a given state *X*_*i*_ must be either 1 or 0. Zero indicates a terminal state.

The **System State** at any point in time is given by the attributes of the individuals (nodes) and contacts (edges) of the network, the simulation time, and the value of user-defined variables.

The values *nodeTrait* and *edgeTrait* are user-defined attributes which may be used to govern interventions (described below). They do not influence the disease transmission or progression. **Interventions** are external modifications of the state of the simulation, where “external” means not governed by the disease model and the contact network itself. An intervention comprises of a trigger and an action ensemble. The action ensemble is only applied if the trigger evaluates to *true*.

The trigger is a function of the systems state and thus may depend on any of the above-mentioned attributes, including the Person Trait DB.

An action ensemble operates on a target set which may contain either nodes or edges. Operations may be performed: (*i*) once per intervention (typically to update variables), (*ii*) for each element within the target set, and (*iii*) for a sampled subset, as well as for the remaining non-sampled elements of the target set. Note that sampling may be nested, i.e., it is possible to sub-sample an existing sample set. Furthermore, it is possible to delay the operation to a later point in the simulation.

Prior to the simulation, the state of any individual *P* must be initialized. Initialization is a special case of an intervention where the trigger is omitted.

**Table V.** EpiHiper state values of nodes and edges

| Object | Value | Access | Description |
| --- | --- | --- | --- |
| system | time | r | the current time (mapped from iteration number/tick) |
| node | id | r | PID of node |
| node | infectivity | rw | infectivity scaling factor |
| node | susceptibility | rw | susceptibility scaling factor |
| node | healthState | rw | health state |
| node | nodeTrait[traitName] | rw | value of nodeTrait[traitName] of node |
| edge | sourceID | r | source vertex ID of edge |
| edge | targetID | r | target vertex ID of edge |
| edge | sourceActivity | r | activity of source of edge |
| edge | targetActivity | r | activity of target of edge |
| edge | active | rw | active flag of edge |
| edge | weight | rw | weight of edge |
| edge | edgeTrait[traitName] | rw | value of edgeTrait[traitName] of edge |
| variable | name | rw | a user-defined numerical variable referenced by name |

## Appendix E

### Bayesian Model Calibration

#### Agent-Based Model Calibration

For a particular state, we model the observed time series of logged reported case counts ***y*** = (*y*_1_, *y*_2_,*…, y*_*t*_) as an additive combination of the computational model *η*(***θ***), i.e., EpiHiper at the best setting for the parameters, a systematic model discrepancy term *δ*, and observation error *E*:

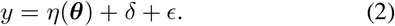

where, ***θ*** denotes the set of parameters that we are interested in finding the plausible values given the observed data. To begin with, a limited number of simulations are run at pre-specified parameter settings, based on which a Gaussian Process (GP) emulator [46] is specified, so that uncertainty in the simulator output at untried setting ***θ***^***\****^ can be accounted for in the analysis. To handle the multivariate nature of the simulation output, as well as the observations, a basis representation is used for *η*():

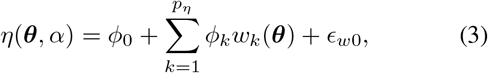

where *p*_*η*_ is the number of basis functions, and *E*_*w*0_ accounts for error due to the limited number of basis functions used. We have used *p*_*η*_ = 5 and eigenvector basis functions (*ϕ*_*k*_) for this analysis. Each individual basis coefficient *w*_*k*_(***θ***) is given a zero mean GP prior having Gaussian covariance functions, with hyper-parameters controlling the marginal variance, the correlation length in each of the component directions of ***θ***, and a nugget term so that interpolation is not necessarily enforced depending on the resulting posterior distribution.

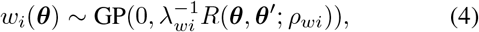

where *λ*_*wi*_ is the marginal precision of the process and the correlation function is given by

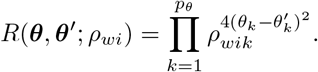

The discrepancy term *δ* is also modeled using a basis representation over time. In this case, the discrepancy basis vectors are 1-d normal kernels with an sd of 15 days; the kernels are spaced 10 days apart.

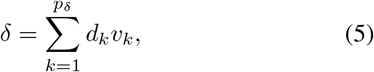

where *p*_*δ*_ = 7 and each *v*_*k*_ has a zero mean normal prior with precision *λ*_*δ*_.

All precision hyper-paremeters *λ* are given suitable gamma priors and the correlation hyper-parameters *ρ* are given beta priors. For calibration parameters ***θ***, we assume a uniform prior defined by their ranges. Gaussianity assumption for the observation error *E* along with prior specifications complete the posterior of ***θ***. Detailed derivation of the likelihood and posterior can be found in [18]. This posterior is explored via MCMC using the GPMSA [23] tool in Matlab.

*Metapopulation Model Calibration* For each county *c* in a state, we model the observed time series of reported case counts 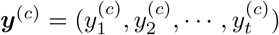 as noisy realization from the underlying metapopulation model *η*(***θ***), with additive Gaussian noise. The noise standard deviation is assumed to be 20% of the daily case counts. Assuming independence between counties, the joint likelihood can be written as the product of *C* multivariate gaussian pdfs:

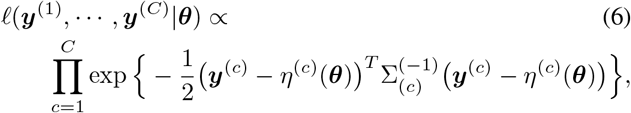

where, *C* is the total number of counties in the state, *η*^(*c*)^(***θ***) represents the simulation output for county *c* at parameter setting ***θ***, and Σ_(*c*)_ contains the error covariances for the same county [17].

Unlike Agent-Based Models, the metapopulation model is cheap to run, hence, calibration is carried out by directly simulating from the model in the Markov Chain Monte Carlo (MCMC) loop. The parameters of interest ***θ*** are given uniform priors based on their ranges. We use metropolis update in the Markov chain. Inferences and predictions are carried out based on samples from the posterior distribution.

## Appendix F

### Case study

#### Case study 2: County-level projections

For our case study on the dynamics of COVID-19 at the state level, we adopted a combination of mechanistic metapopulation and agent-based modeling frameworks similar to the US national-scale models we have employed for forecasting spatio-temporal spread of seasonal influenza. Our model represents SEIR disease dynamics across counties. The overall workflow is shown in Figure 5. The disease dynamics were modified to reflect the transmissivity of asymptomatic and pre-symptomatic COVID-19 patients, as well as the disease parameters calculated from early COVID-19 estimates. We expect these disease characteristics to be refined over time. Additional data sources we rely upon to train our model include: **(a)** County-level confirmed case datasets (initial conditions) queried from the COVID-19 Surveillance Dashboard, an online web application that provides a history of curated COVID-19 incidence data. **(b)** Counts by date of illness onset and other outcomes, such as hospitalizations and ICU treatment, obtained from the state health department; **(c)** Hospital bed and ventilator counts obtained from individual hospitals, as well as from the 2018 American Hospital Association (AHA) estimates.

We model five different scenarios. One is the worst-case scenario, where limited social distancing is observed. The remaining four assume a start date of March 15, 2020 for intense social distancing, and are further differentiated by the proposed end date for intense social distancing (April 30, 2020 and June 10, 2020) and reduced transmissibility rates (25% and 50%).

Transmissibility and infectious duration parameters are calibrated based on county-level confirmed cases, utilizing a Bayesian model calibration approach. Logged values of cumulative counts were modeled as noisy realization of the underlying disease dynamics, with noise following a multiviate Gaussian distribution. The full posterior was obtained using this likelihood and uniform prior specification on disease parameters. Markov Chain Monte Carlo was used to explore the posterior distribution, and uncertainty bounds were obtained based on the samples from the posterior.

#### Case study 3: Calibrating agent-based model

In this section we demonstrate the calibration-prediction workflow described earlier using Virginia as an example. We have done the same for all 50 states and Washington DC for many cycles. In this case study, we started with cumulative confirmed case counts of Virginia through April 11, 2020. We took the “best guess” disease model defined in [8], and vary two parameters: the disease transmissibility and the ratio between symptomatic and asymptomatic cases. We implemented a school closure (SC) mitigation, which started on March 16, 2020, and would extend through the end of August; and a stay-at-home order (SH), which started March 31, 2020, and would expire on June 10, 2020. The timing of these mitigations is real. We also implemented voluntary home isolation (VHI) of symptomatic cases (VHI). We vary compliance rates on SH and VHI mitigations, and assume 100% compliance on SC. With SC, all schools, including colleges, are closed so *school* type contacts are disabled. With SH, we disable all contacts of compliant people except their *home* type contacts (with their family). With VHI, compliant people stay at home and their non-*home* type contacts are disabled.

We created a design of 100 configurations (prior) with the Latin hypercube sampling method [35]. After simulating these configurations and aggregating the output, we ran the Bayesian calibration to obtain another 100 configurations (posterior). Figure 15 shows the comparison of the prior and posterior configurations. Figure 16 shows the visualization of the calibration that helps us to decide the goodness-of-fit. After simulating the posterior configurations, we were able to make a prediction – Figure 17 shows our forecast for the next eight weeks for cumulative confirmed case counts.

**Figure 16.**
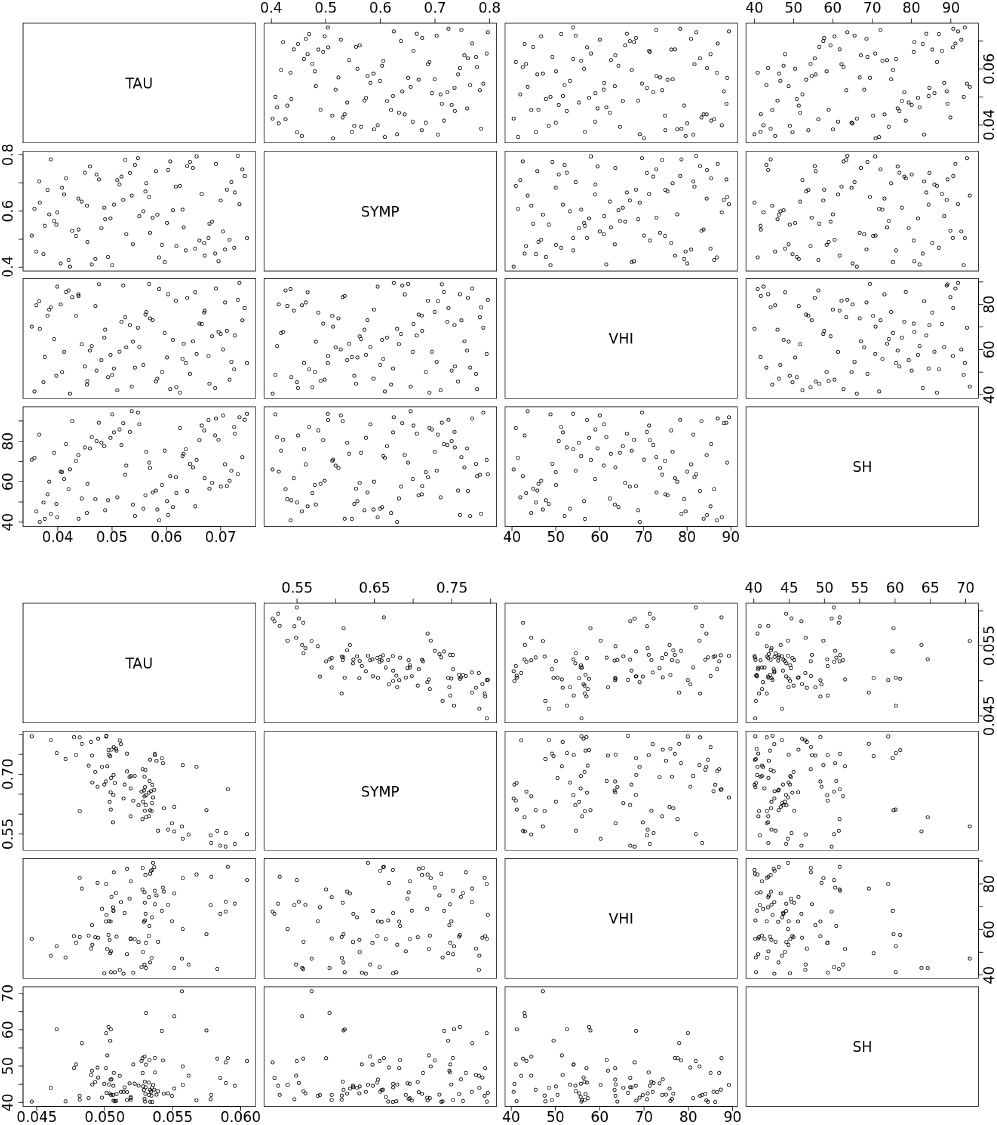
Visualization of calibration produced by the GPMSA tool is used to evaluate the results. We mainly consider the left panel plot, where the blue marks show the ground truth and the green curves show the 95% uncertainty interval of the emulated data from the GP emulator. The result is good if the ground truth falls between the green curves. Otherwise, we may need to continue calibrating with more iterations.

**Figure 15.**
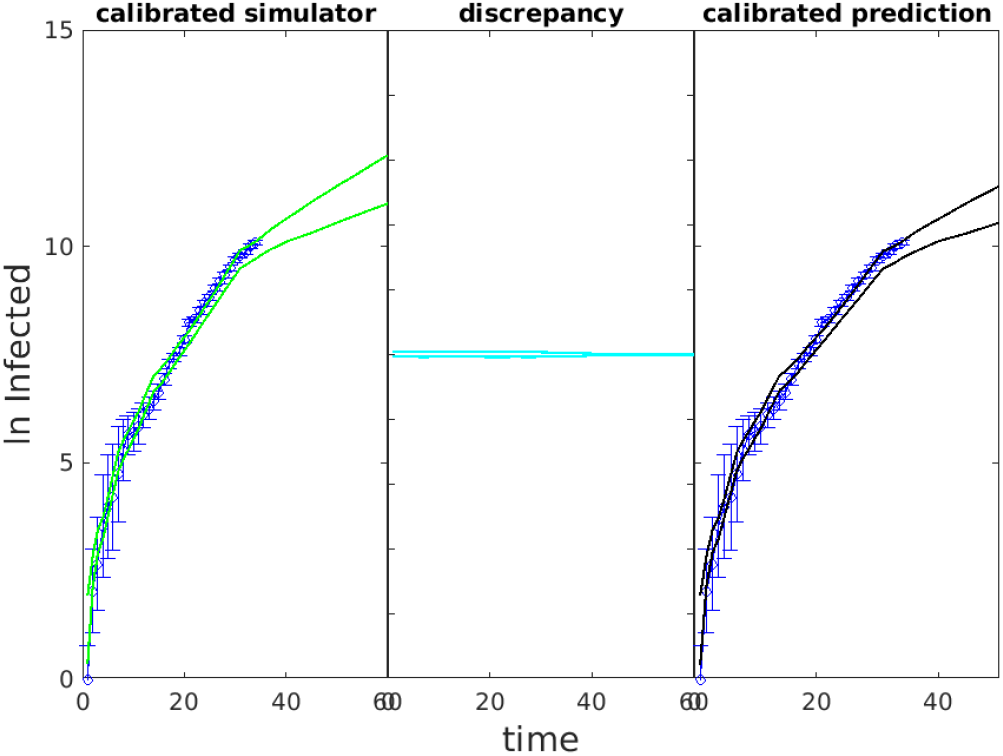
Scatter plots of varying parameters in the prior (top) and posterior (bottom) designs show how the distributions are changed by the calibration. In the prior design, the parameters are evenly distributed in the pre-specified space. After calibration, transmissibility (TAU) and symptomatic fraction (SYMP) seem to be negatively correlated and both distributions are tightened; SH compliance distribution is concentrated towards lower values; VHI compliance distribution seems unchanged.

**Figure 17.**
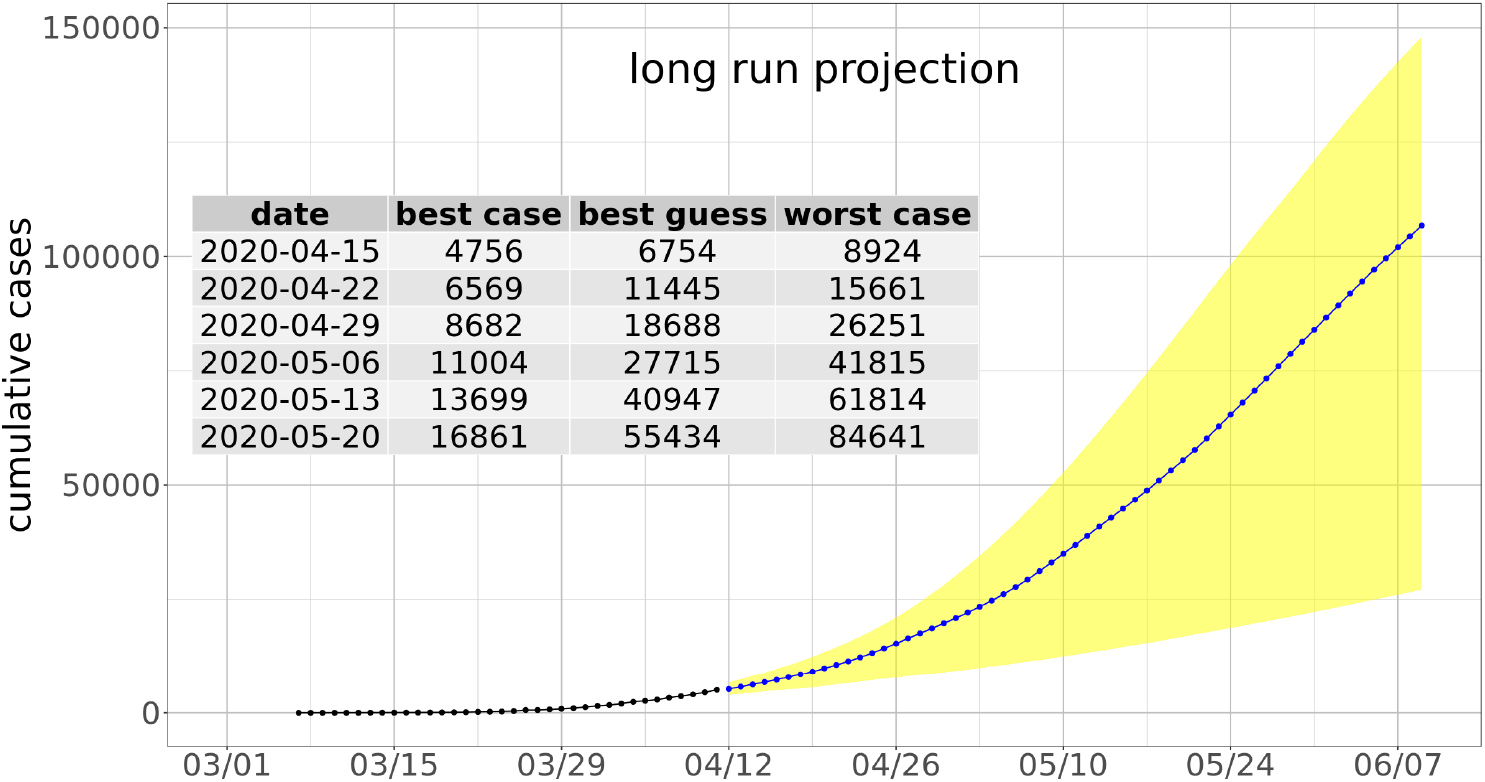
Prediction of the cumulative confirmed case count for the state of Virginia for each day of the two months after April 11, 2020. The black curve represents the reported case counts up to April 11, while the blue curve is the median of prediction and the yellow band shows the 95% uncertainty interval.

## Footnotes

1 This is an extended version of the paper with the same title that was published at the 35th IEEE International Parallel and Distributed Processing Symposium.

2 More details on how our models are used can be found at: https://www.vdh.virginia.gov/coronavirus/category/covid-19/model

3 More information about our work can be found at https://biocomplexity.virginia.edu/project/covid-19-pandemic-response

4 https://www.postgresql.org/

5 https://github.com/nytimes/covid-19-data

6 https://nssac.bii.virginia.edu/covid-19/dashboard/

7 https://coronavirus.jhu.edu/map.html

8 https://www.globus.org/

9 https://github.com/ihmeuw/covid-model-seiir-pipeline

10 https://covid-19.bsvgateway.org/

11 https://reichlab.io/

12 https://landscan.ornl.gov/.

13 http://www.openstreetmap.org

14 https://sedac.ciesin.columbia.edu/data/collection/gpw-v4

